# Comprehensive molecular characterization of cutaneous squamous cell carcinoma reveals determinants of metastatic progression

**DOI:** 10.64898/2026.07.17.26358051

**Authors:** Barbara Rentroia-Pacheco, Harsh Sharma, Lara Pozza, Joleen J.H. Traets, Bishal Tandukar, Olivia F.M. Steijlen, Romy Ruiter, Noel Cruz-Pacheco, Daphne Huigh, Arne Van Hoeck, Yan-Ting Chen, Beatriz Infante, Defne Baskurt, Vignesh Arunachalam, Celeste J Eggermont, Amanda Bas-Cristóbal Menéndez, Tamar Nijsten, Harmen J. G. van de Werken, Antien L Mooyaart, Domenico Bellomo, Marlies Wakkee, A. Hunter Shain, Loes M. Hollestein

## Abstract

Cutaneous squamous cell carcinoma (cSCC) is the second most common form of cancer worldwide. While most cSCCs are not life-threatening, 2-5% of patients develop metastases. To better understand what causes some cSCCs to progress to metastatic disease, we assembled a nationwide cohort of 19,120 patients with clinico-pathologically annotated tumors linked to metastatic outcome. RNA-sequencing was performed on 378 tumors, and whole-exome sequencing on 147, with balanced numbers of tumors that progressed to metastatic disease (cases) and did not (controls). UV radiation was the dominant mutational signature with additional contributions from aging, APOBEC activity, and, in immunosuppressed patients, azathioprine exposure. We identified 38 genes under selection across a core set of signaling pathways. Gene expression clusters were primarily associated with the differentiation state of tumor cells and secondarily with the composition of the tumor microenvironment. Several mutational and transcriptional programs were associated with metastasis, including a dedifferentiated gene expression signature, activating mutations in the RAS signaling pathway, loss-of-function alterations in the SWI/SNF chromatin remodeling complex, and specific arm-level copy number alterations. A 23-gene expression signature was built to predict metastasis from primary cSCC tissue. The signature was validated in two independent cohorts (N=102 and 52), where it predicted metastasis independently of staging systems. Together, these findings provide the most detailed molecular portrait of cSCC to date and establish an assay for risk stratification suitable for clinical implementation.

## Introduction

Cutaneous Squamous Cell Carcinoma (cSCC) is a type of skin cancer that is the second most common form of cancer worldwide^1,2^. Given its high and rising incidence, cSCC poses a major public health burden^3–5^. However, it has been inaccurately perceived as a non-life-threatening cancer, and consequently understudied. Many national cancer registries exclude cSCC from registration^6,7^ and large-scale molecular atlases, such as The Cancer Genome Atlas (TCGA), have not profiled cSCC. Consequently, there is a lack of high-quality epidemiological studies and large-scale biomarker discovery studies describing cSCC, which has limited our ability to understand and effectively treat the disease.

The perception of cSCC as a non-life-threatening cancer stems from the relatively low risk of metastasis from primary cSCCs (2–5%)^8,9^ compared to other cancers. However, while an individual tumor may pose low risk to a patient, since cSCC is so common, the total morbidity and mortality across the entire population is high. Moreover, managing cSCC remains challenging. Some treatments, such as extensive surgeries and/or (adjuvant) immuno- or radio-therapy, require careful consideration because many patients are elderly, frail, or immunosuppressed, as these interventions may be disfiguring and carry substantial morbidity. When tallying the cumulative costs on our healthcare system, cSCC is arguably worse than melanoma. It kills a similar number of patients while affecting many more, thus burdening healthcare systems with billions of dollars in management costs on top of its death toll^5^, much of which could be avoided with better prognostic stratifications.

Most clinicians use staging systems developed by Brigham and Women’s Hospital (BWH)^10^ or the American Joint Committee on Cancer (AJCC)^11^, however, these are insufficient predictors of metastatic risk^12^. We recently developed a new system (the Erasmus MC model) based solely on clinical and histopathologic features that outperforms the BWH and AJCC systems^13^, but there remains limitations to clinical/pathologic staging systems. For instance, these systems require complete pathological assessment of the excised tumor, rendering them unsuitable for use with biopsy material and limiting their utility in pre-operative decision-making. Furthermore, observer variability in pathological assessment limits reproducibility across centers^14^.

As such, better biomarkers are needed to quantify metastatic risk in patients with primary cSCC. This knowledge could guide treatment strategies, indicating which patients may require margin-controlled surgery, additional imaging and/or adjuvant therapies, versus those who only need limited follow-up screening. To this end, gene expression signatures to predict cSCC metastasis have been developed^15–17^. Despite their potential, these signatures were developed in cohorts that do not reflect the actual distribution of cases across T-stages and have a low number of metastatic events. In addition, external validation in representative cohorts is still lacking.

To address these gaps in knowledge, we established the D-SQUAME study, one of the World’s largest and most deeply annotated datasets of cSCC^18^. Next, we performed a large-scale genomic and transcriptomic analysis of cSCCs drawn from this study (Fig. 1A). From these discovery analyses, we identified mutational and expression markers predictive of metastatic outcome. More generally, because cSCC was left out of large-scale sequencing studies, we envision the data here will serve as an important resource for the genomics community.

**Figure 1.**
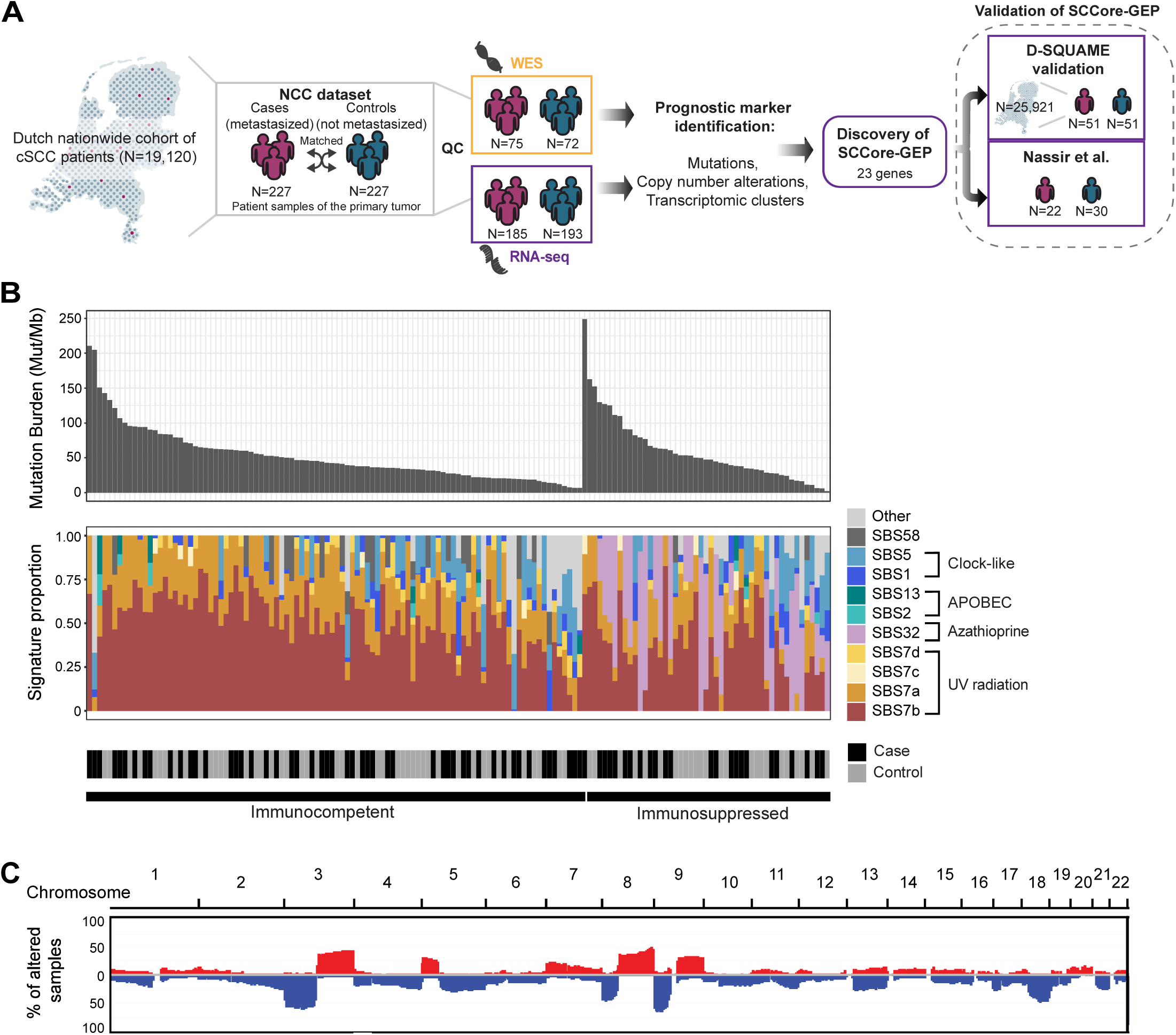
Mutational landscape of cutaneous squamous cell carcinoma. **A.** Overview of cohorts used in this study. NCC: Nested case-control **B.** Top: Mutation burden of each tumor (mutations per megabase). Bottom: Proportion of mutations assigned to each COSMIC mutational signature in each tumor. Immunocompetent indicates tumors from patients without any known immunosuppressive condition. Immunosuppressed indicates tumors from patients who received an organ transplant or had a hematological malignancy before cSCC diagnosis (see methods). **C.** Genome-wide chromosomal alterations (red = gain, blue = deletion).

## Results

### Cohort description

To facilitate molecular studies, we first assembled the D-SQUAME study^18^ – a nationwide cohort of cSCCs with detailed clinical/histopathologic information and long-term outcomes. We integrated data from the Netherlands Cancer Registry (NCR), the Dutch Nationwide Pathology Databank (Palga), and the Netherlands Organ Transplant Registry. The organ transplant registry was used to identify solid-organ transplant recipients, and the NCR to identify patients with hematological malignancies – these patients show an increased risk of cutaneous squamous cell carcinoma due to immunosuppression^2,19^. We identified all 19,120 patients diagnosed with a first cSCC between 2007 and 2009, selecting this interval to ensure adequate follow-up to distinguish lesions that progressed to metastatic disease from those that did not. From this cohort, we performed RNA-sequencing on 378 tumors and an additional round of whole-exome DNA sequencing on 147 tumors (Table S1, Fig. S1A). All patients were treatment-naïve and free of metastatic disease at the time their primary tumors were diagnosed.

We used a Nested Case-Control (NCC) study design to select balanced numbers of tumors that progressed to metastatic disease (cases) and did not (controls). A nested case-control study is a cost-effective way to sample a meaningful number of events (in this case, tumors that progress to metastatic disease) while retaining the ability to extrapolate absolute risk estimates to the whole population by weighting each sample according to how representative it is of the source cohort^20^.

As expected, >80% of cSCCs in the nationwide cohort of 19,120 tumors were early-stage, with an extremely low metastatic risk (Fig. S1B). Therefore, when selecting tumors for molecular analyses, we intentionally selected controls whose clinical and histopathologic features more closely resembled the cases (Fig. S1B, C). This was accomplished by pairing each case with a control from the same pathology lab that had a similar clinical-pathologic risk score^13^. Our strategy increased the likelihood that any molecular biomarker predictive of metastatic disease would provide prognostic information that complements, rather than simply recapitulates, established staging systems.

### Landscape of genetic alterations in cSCC

Studying the mutational landscape of cSCC is difficult because these tumors tend to have low neoplastic cellularity, owing to dense immune infiltrates and reactive hyperplasia mixed with neoplastic cells^21^. An additional challenge is that most tumors were derived from formalin-fixed, paraffin-embedded (FFPE) blocks that were more than 10 years old. To overcome these obstacles, we applied rigorous quality control standards to ensure that we were statistically powered to detect clonal mutations in each tumor (see methods). To our knowledge, no prior genomic study of cSCC has analyzed a cohort of this size while meeting comparable quality control criteria.

cSCCs had high mutation burdens (median of 44 mutations/megabase) (Fig. 1B). UV radiation was the dominant mutational signature with additional contributions from aging, APOBEC activity, and, in immunosuppressed patients, azathioprine exposure^22^ (Fig. 1B). Recurrent copy number alterations were observed, including gains of 3q, 5p, 8q and 9q as well as losses of 3p, 8p, 9p, and 18q (Fig. 1C, S2A-D). There were no statistically significant differences in the total mutation burden or fraction of the genome altered of cases versus controls or between tumors from immunocompetent versus immunosuppressed patients (Fig. S2E-F).

Due to its high mutation burden, it is challenging to discriminate driver mutations from passenger mutations in cSCC^23^. To maximize statistical power, we combined data from the 147 tumors sequenced here and 83 tumors from a previous meta-analysis of cutaneous squamous cell carcinoma^21^ and used IntOGen^24^ to identify genes under selection (Table S2). Mutations were also checked for overlap with the OncoKB^25^ database to detect drivers that were too rare in this dataset, alone, to show patterns of selection but are known to be under selection in other cancers (Table S3). Furthermore, we annotated genes with focal amplifications and deep deletions.

In total, we identified 38 driver genes, converging on a core set of signaling pathways (Fig. 2A, S3). Most tumors harbored alterations inactivating the Notch, p53, Hippo, and Rb pathways. Tumors also frequently carried loss-of-function mutations in *KMT2D* (53.7%), a key chromatin regulator of epidermal differentiation^26^, and gain-of-function mutations in the *TERT* promoter (58.5%). To a lesser extent, they had alterations disrupting the SWI/SNF chromatin remodeling complex (45.6%), activating the RTK–RAS–MAPK/PI3K pathway (32.0%), or inactivating TGF-β signaling (22.4%). TGF-β pathway mutations were now significant when compared to the previous meta-analysis of cSCC^21^, though Cammareri and colleagues had previously noted mutations in this pathway and provided functional evidence that it was tumor suppressive^27^. We also refined the specific genes disrupting other pathways already known to drive cSCC. Additional low-frequency driver genes are likely to exist, but this study represents, to our knowledge, the largest statistically powered cancer gene discovery effort to date in this tumor subtype.

**Figure 2.**
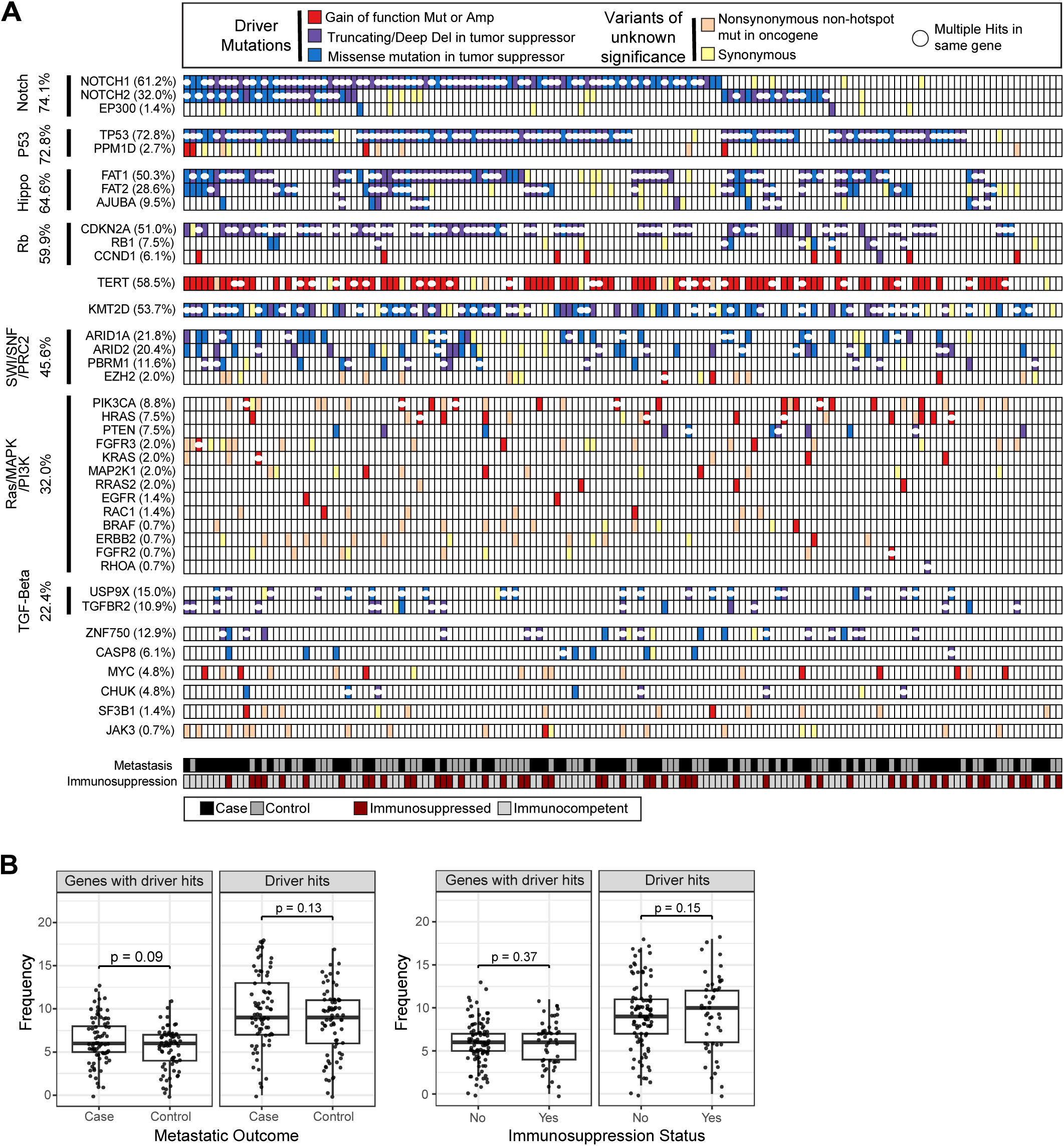
Landscape of driver mutations in cutaneous squamous cell carcinoma. **A.** Tiling plot of genetic alterations (rows) in each tumor (columns). Genes with at least one driver mutation are shown. Genes are grouped into pathways and ordered by pathway mutation frequency (highest to lowest). White circles indicate the gene has multiple alterations within the same tumor - when this occurs, the tile is colored to indicate the most consequential alteration. For the computation of the percentage of sample with driver alterations in each gene or pathway, only driver mutations are counted. **B**. Number of genes with driver hits and number of total driver hits per tumor, stratified by metastatic outcome and immunosuppression status. Differences between groups were assessed using linear regression adjusted for callable coverage.

Tumor suppressor genes classically acquire two “hits” to eliminate the activity of both parental alleles^28,29^. We used copy number data and mutant allele frequencies to infer genes with bi-allelic alterations (see methods, Fig. S4). While IntOGen did not consider the number of hits when identifying genes under selection, most tumor suppressor genes had alterations affecting both alleles in the majority of the samples (Fig. S4A), further supporting their case as driver genes. A major exception was SWI/SNF genes (*ARID2* and *ARID1A*), for which selection is more complex. SWI/SNF mutations alter, but do not fully ablate, chromatin remodeling activity, and hence bi-allelic mutations are sometimes not tolerated^30^. Several oncogenes, notably *PIK3CA*, also showed an increase in dosage of the oncogenic allele (Fig. S4B). While acknowledging the caveat that we are missing low-frequency pathogenic alterations, the median cutaneous squamous cell carcinoma is driven by 9 hits across 6 genes (Fig. 2B). We also tested whether mutations in driver gene pairs co-occurred or were mutually exclusive; however, no gene pair showed a significant association after multiple hypotheses testing (Table S4).

A major strength of this study is the ability to link genetic alterations with metastatic outcomes. Hazard ratios were calculated to compare metastatic event rates between tumors with and without mutations in individual genes, signaling pathways, or arm-level copy number alterations (Fig. 3A–C). We then applied a regularized Cox regression model to identify combinations of features that best predicted metastatic risk across bootstrap resamples (Fig. 3D, S5A). Bootstrapping was used to evaluate the stability of feature selection and to mitigate overfitting. Across bootstrap iterations, model performance was consistent in out-of-bag samples and effectively stratified tumors into risk groups (Fig. S5B-C). Poor prognosis was associated with mutations in the SWI/SNF, RTK–RAS–MAPK/PI3K, and TGF-β pathways; *MYC* gain (chr8q) or mutation; losses of 11q, 8p, or 3p; and mutations in *CASP8*. In contrast, a favorable prognosis was associated with *KMT2D* mutations, gain of 5p or loss of 4q or 10q.

**Figure 3.**
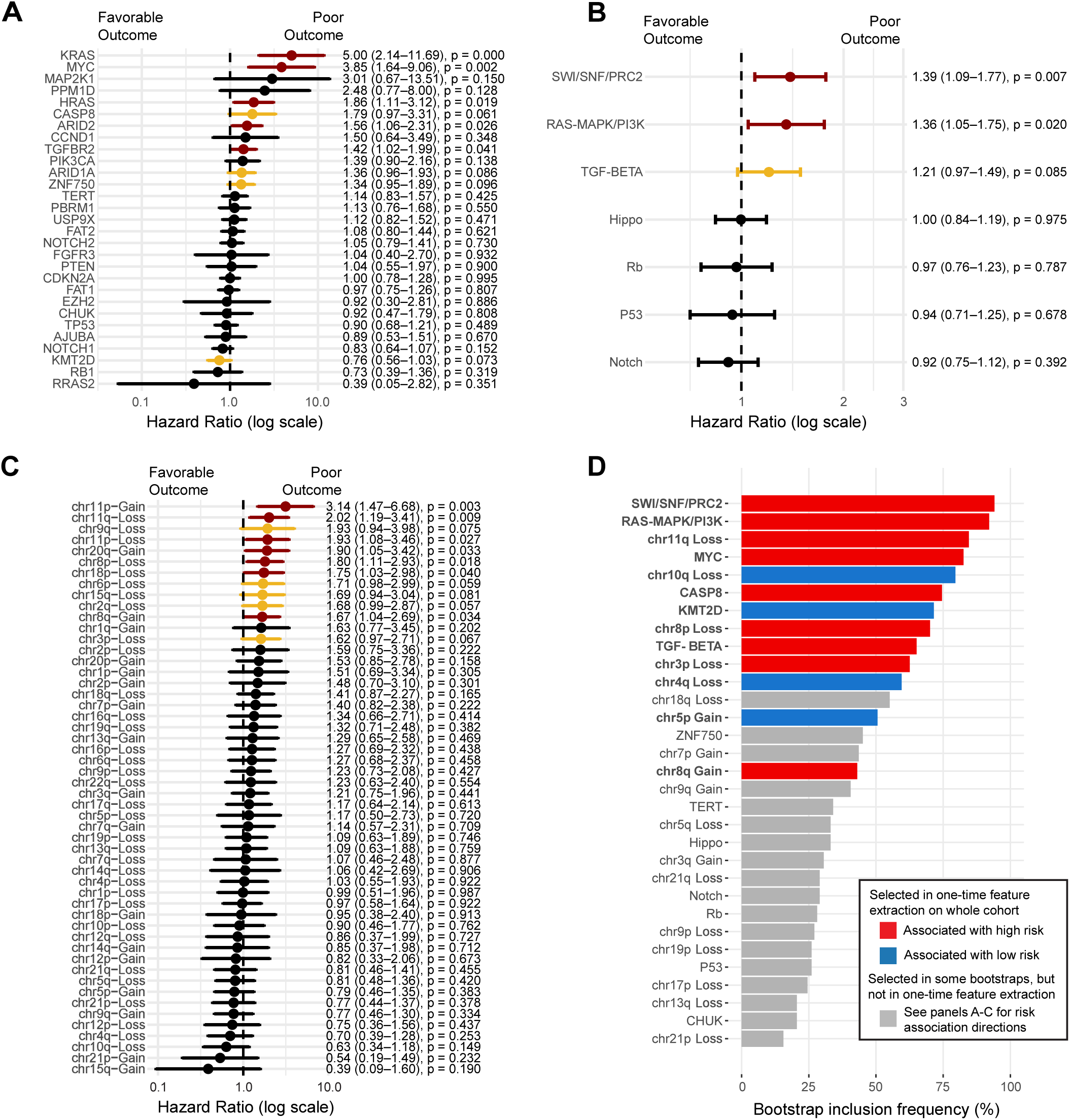
Genomic alterations associated with metastatic outcome. **A.** Association between driver mutations and metastatic outcome for all genes with driver mutations present in at least 3 samples. Association was assessed using a Cox proportional hazards regression model, with gene hit count as the independent variable and sample callable coverage included as an adjustment covariate. Hazard ratios, associated 95% confidence intervals and Wald p-values for gene hit count are shown. p-values < 0.1 are colored in yellow, and p-values <0.05 are colored in red. **B.** Genes with driver mutations were grouped into pathways and association between the sum of all hits in a pathway and metastatic outcome was computed, with adjustment for sample callable coverage. **C**. Association between chromosomal alterations and metastatic outcome, adjusted for sample callable coverage. Only chromosomal alterations present in at least 10 samples are shown. **D**. The most predictive combination of pathway and copy number alterations was selected using a multivariable regularized Cox model. The selected variables (in bold) were frequently selected by regularized Cox models across bootstrap samples (N=200).

We also computed odds ratios to determine whether specific genetic alterations were enriched in tumors from immunocompetent versus immunosuppressed patients. Mutations in the RTK-RAS-MAPK/PI3K pathway were more common in tumors from immunosuppressed patients, though the association just missed statistical significance (Fig. S6).

### Gene expression landscape in cSCC

Next, we clustered the RNA-sequencing data. The tumors were a mixture of surgical excisions and biopsies, and whole-slide RNA-sequencing, without macrodissection, was performed on each sample. To avoid batch effects due to the different composition of tissues from excisions versus biopsies, we performed unsupervised hierarchical clustering on the most variable genes within excisions and used the biopsies to validate the sample/gene clusters.

Consensus clustering identified three sample clusters and six gene expression clusters (Fig. 4A, Table S5). These clusters were reproduced in the biopsies (Fig. S7A-C). Samples clustered into groups with favorable, intermediate, and poor metastatic outcomes (Fig. 4B). Pathway overrepresentation analysis showed that gene expression clusters were largely driven by epithelial differentiation and were strongly associated with histopathologic differentiation (p<0.001). Differentiated tumors had (terminal) squamous cell differentiation programs with the longest metastasis-free survival. Basal-like tumors showed a higher expression of basal squamous cell programs with an intermediate metastatic outcome. Finally, Mesenchymal-like tumors exhibited poorly differentiated programs with the shortest metastasis-free survival. Further, there was a shift in metabolic signatures and proliferation. Differentiated tumors were enriched for lipid metabolic programs, and Mesenchymal-like tumors preferentially expressed the glycolytic pathway, consistent with the Warburg effect. They also showed higher expression of genes involved in the cell cycle. Our results support the model that tumor differentiation, metabolic and proliferation status are associated with time to metastasis.

**Figure 4.**
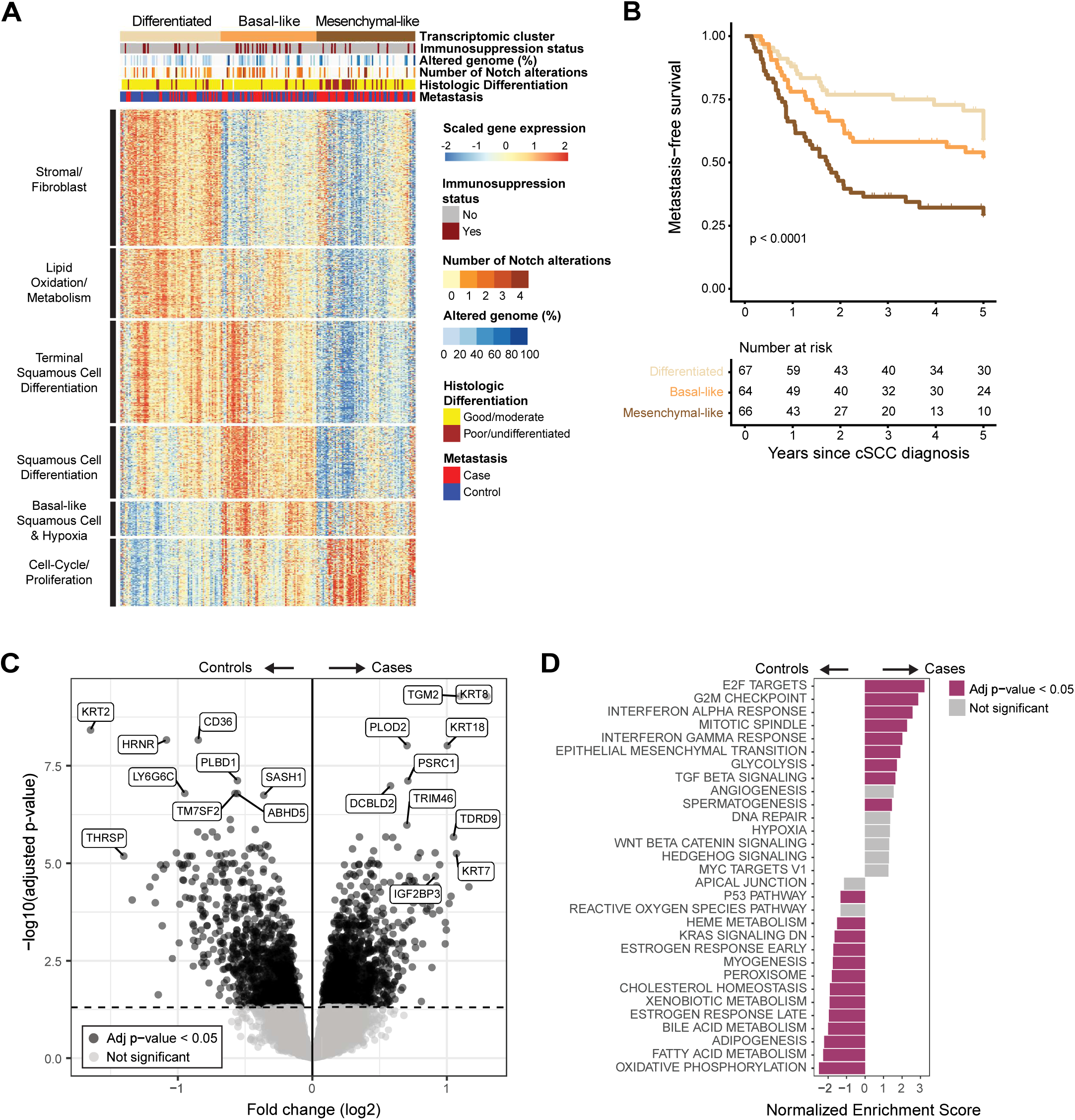
Landscape of gene expression patterns in cutaneous squamous cell carcinoma. **A.** Clustering of cSCC excision samples (N=197). Within each cluster, samples are ordered according to hierarchical clustering using Spearman’s correlation and average linkage. Rows represent the 1000 genes with the highest association to the sample clusters. The tile plot above the heatmap shows the distribution of pathological characteristics and genomic alterations that were found to be significantly associated with the clusters (see Table S6), tiles with missing values are colored in white. **B**. Kaplan-Meier curve of the three cluster groups. Follow-up truncated at 5 years. **C.** Volcano plot showing differentially expressed genes between metastasizing and non-metastasizing CSCC, adjusted for sample type (excision vs biopsy). Significant genes were defined as those with an adjusted p-value < 0.05, indicated by a dashed horizontal line. **D.** Gene Set Enrichment Analysis (GSEA) showing the top 15 upregulated and top 15 downregulated Hallmark gene sets in metastasizing versus non-metastasizing CSCC, adjusted for sample type.

Tumor microenvironment signatures were also apparent. Favorable outcome tumors preferentially expressed stromal/fibroblast-rich gene expression programs, whereas intermediate outcome tumors were enriched for hypoxia-related signatures. Curiously, immune-related programs did not drive the unsupervised clustering. However, immune signatures were evident when the data were analyzed in a targeted manner. For example, tumors from immunocompetent patients showed enrichment of interferon-α and γ signaling and other immune-related pathways A-B). However, these immune signals were outweighed by differentiation- and metabolism-associated programs in unsupervised analyses. Prior studies have reported immune composition during progression from actinic keratoses to primary cutaneous squamous cell carcinoma^31,32^. If immune evasion is, indeed, an early event in skin carcinogenesis, it would already be present across most tumors in this cohort and likely overshadowed by other biological processes, such as differentiation.

We additionally tested for associations between sample clusters and clinical/genomic features (Table S6 and Fig. 4A, top tiles). Several pathological risk factors, including BWH and AJCC8 stages, increased from the differentiated to the mesenchymal tumors (p<0.001) as well as the proportion of genome altered (p=0.0015). Curiously, Basal-like tumors were associated with a higher percentage of Notch pathway mutations (p=0.029).

We also performed supervised differential gene expression analysis comparing cases and controls, identifying 2,300 genes that differed between the two groups (Fig. 4C). The transcriptional programs evident in this supervised analysis mirrored those identified by unsupervised clustering (Fig. 4D). A previous study profiled the gene expression landscape of earlier phases of skin carcinogenesis through comparison of normal skin, actinic keratoses, and primary cutaneous squamous cell carcinomas^31^. They defined a differentiation–progenitor–like (DvP) signature that also overlapped with our genes (Fig. S7D), confirming that differentiation is a dominant signal distinguishing cases and controls.

To distill these gene expression programs into a prognostic signature, amenable to clinical deployment, we implemented a regularized Cox regression model to select the combinations of genes whose expression is best able to discriminate cases from controls. As model input features, we tested the previously defined DvP genes (biology-inspired genes), genes defined exclusively in this study (data-driven genes), and integrations of the two gene sets (SFig. 9A). The entire procedure, including feature filtering and selection, was internally validated through bootstrapping (SFig. 9B) to get a reliable model performance estimate. The models based on DvP genes or exclusively on our genes performed well (SFig. 10A), but the model that combined features from both sets, with a late feature integration approach, achieved a high performance with fewer input genes, showing an internally validated weighted C-index of 0.75 (95%CI 0.66-0.80) (SFig. 10B). The final model, obtained by running the same pipeline on the entire dataset, yielded a 23-gene signature (Table S7), SCCore-GEP (Squamous Cell Carcinoma Outcome Risk Estimation Gene Expression Profile).

Out of the 23 SCCore-GEP genes, 10 were upregulated in cases and were associated with cell migration and stemness. The remaining 13 genes were upregulated in controls and were mostly associated with epidermal differentiation (Fig. 5A, left). We performed spatial transcriptomics on a subset of tumors to determine where the gene expression of the SCCore-GEP genes derived from (Fig. 5A, right). Most genes were primarily expressed in tumor keratinocytes and secondarily in normal epithelial cells or immune cells. *PLA2G2A* gene was an exception, as it was expressed mostly in fibroblasts.

**Figure 5.**
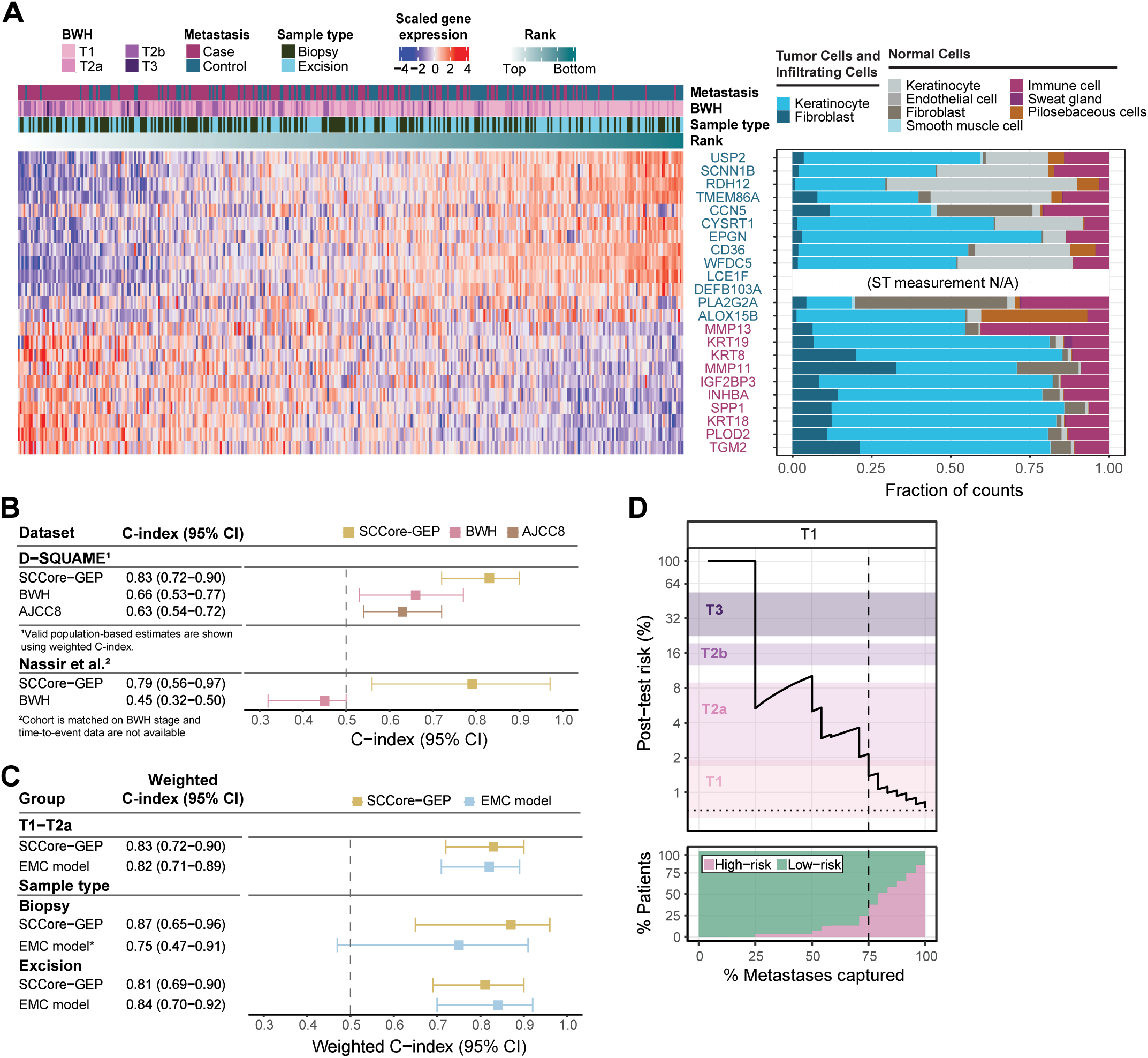
SCCore-GEP: characterization of gene expression and validation performance. **A.** Scaled gene expression values of the 23 SCCore-GEP genes in the D-SQUAME discovery dataset. Samples are ordered according to the ranked SCCore-GEP risk output. Metastasis status, BWH stages, and sample type are indicated by colour annotations above the columns. Spatial transcriptomics on 4 case-control pairs (N=8 samples) from the discovery dataset reveals the read distribution of each of the 23 genes across cellular compartments. **B**. Assessment of the discriminative performance for metastasis prediction of the SCCore-GEP, BWH, and AJCC8 staging systems in T1-T2a tumors of the D-SQUAME validation and the Nassir et al datasets. **C.** Assessment of the discriminative performance for metastasis prediction of the SCCore-GEP, and EMC model in T1-T2a tumors of the D-SQUAME validation dataset with and without sample type stratification. Performances are reported in terms of weighted C-index and 95% confidence intervals (see methods). *Whenever an excision sample was available, EMC model predictions were computed using pathological variables scored on the excision sample. **D.** SCCore-GEP post-test risk probability across different risk thresholds. Varying the risk threshold on the predicted SCCore-GEP risk results in different stratifications of the patients into a high-risk and a low-risk groups. We show the post-test risk (i.e., Positive Predictive Value/precision) versus the percentage of metastases captured (i.e., sensitivity/recall) in the high-risk group, for different risk thresholds, within BWH T1 patients (N samples in NCC dataset= 62). Color bands correspond to the 95% confidence intervals of the nodal metastasis prevalence in each BWH stage, as reported in Zakhem et al. Horizontal dotted lines indicate the pre-test risk in BWH T1 patients in the D-SQUAME validation dataset (0.70%). The lower panel shows the percentage of post-test low-risk and high-risk patients versus the percentage of metastases captured (sensitivity/recall). The interval from 0 to the dashed vertical line corresponds to risk thresholds for a potential upstaging of the patients.

### Validation of a molecular signature to predict metastatic outcome in patients with primary cSCC

Mutational and gene expression features were both prognostic in the discovery cohort, but they were identified from datasets with incomplete overlap, making it difficult to directly compare their performance. To determine whether to pursue a prognostic biomarker based on mutations, gene expression markers, or a combination of both, we directly compared their performance on the subset of tumors with available DNA- and RNA-sequencing data (N=126 tumors). We developed (regularized) Cox regression models for each feature set and evaluated their performance through bootstrapping (SFig. 11). Within this sample subset, the combined model and gene expression-only model performed similarly, and both outperformed the mutation-only model. Considering this, for simplicity, we prioritized validation of the gene expression marker set, SCCore-GEP.

SCCore-GEP was validated in two independent cohorts: a more recent set of tumors from the D-SQUAME study, and an external RNA-sequencing cohort made available by Nassir and colleagues^15^ (Fig. 1A, SFig. 12A). Due to the scarcity of T2b samples (N=2 in the D-SQUAME study), validation was focused on BWH T1-T2a patients. SCCore-GEP showed high discriminative performance and outperformed BWH and AJCC8 staging systems in prediction of metastasis in both cohorts (Fig. 5B). In the D-SQUAME validation cohort, controls were sampled randomly to facilitate a population-based comparison with staging systems and the EMC model. SCCore-GEP achieved a weighted C-index of 0.83 (95%CI: 0.72−0.90), which corresponds to an increase of over 0.15 compared to the BWH and AJCC8 staging systems. Similarly, in the Nassir cohort, SCCore-GEP achieved an AUC of 0.79 (95%CI: 0.56−0.97), 0.34 higher than the BWH staging performance. The signature also showed robust performance across sample types (biopsy or excision, Fig. 5C) and discriminated equally well within BWH stages (SFig. 12B).

Compared to the clinicopathological EMC model, SCCore-GEP showed similar performance in all sample groups, except for samples from biopsy material, where SCCore-GEP was 0.12 higher (Fig. 5C). While the performance of SCCore-GEP, as measured by a C-index, was comparable to the EMC model for many subgroups, multivariable analysis showed that SCCore-GEP provides independent predictive value from staging systems and the EMC model (p-value <0.001, SFig. 12C). For completeness, signature performances for all available T-stages and validation cohorts are respectively reported in SFig. 13A-C and 14.

There is no consensus among clinicians on a post-test risk cutoff for patient upstaging^13^; however, we investigated the signature’s potential to identify patients at high-risk of metastasis within traditionally low-risk groups (BWH T1 and T2a) in the D-SQUAME validation cohort (Fig. 5D, SFig. 12D). Within T1 patients, the SCCore-GEP can identify candidates for upstaging – i.e. patients whose true metastatic risk is higher than the metastatic risk in T1 patients reported in the literature^33^. Depending on the desired post-test risk for upstaging, this high-risk group can represent up to 25% of T1 patients, while accounting for up to 75% of metastatic events in this population (dashed line in Figure 5D). SCCore-GEP can also identify up to a sixth of T2a patients with post-test risk compatible with upstaging: depending on the exact cutoff, this group can account for up to two-thirds of the metastatic events (SFig. 12D). Similar high-risk groups can be found within AJCC8 low-risk patients (T1-T2) (SFig. 14).

## Conclusions

Given the enormous burden on public health systems, cSCC is an ideal example of a cancer subtype that would benefit from biomarkers to predict outcomes, thus reducing the burden of over-and under-treatment. However, it is difficult to identify biomarkers without first establishing large-scale clinical and molecular databases from which discovery analyses can be performed. Here, we generated, to our knowledge, the world’s largest and most comprehensive resource of cSCC by integrating national clinical registries and multi-omics profiling of rigorously selected tumors with known metastatic outcomes.

Our analysis of whole-exome sequencing and RNA-sequencing data encompasses more than twice as many tumors as the next largest studies^21,34^. Prior cancer gene discovery analyses were underpowered to identify driver genes in cSCC^23^, but this study provides the clearest portrait of genes under selection in cSCC to date. We did not observe genetic subtypes of cSCC, but some cSCCs are genetically more advanced, with mutations in the SWI/SNF, Ras, and TGF-beta pathways, and these tumors are more likely to progress to metastatic disease. The Pan-Cancer Analysis Working Group (PCAWG) estimates that most cancers are driven by 4-5 driver genes^35^, but cSCCs are driven by 9 hits spread across 6 genes, mostly affecting tumor suppressors. As the first line of defense against the environment, skin epithelial cells likely evolved mechanisms to avoid simple routes to transformation from one or a few powerful mutations.

Given the evidence of high immunogenicity in cSCC, it was somewhat surprising that immune gene expression programs did not drive unsupervised clustering. The tumors in the discovery cohort were mostly collected between 2007 and 2009, capturing salient metastatic biology without selective pressures introduced by drug treatments such as immunotherapy. A future case–control study stratified by response to immunotherapy would be expected to reveal stronger immune signals, but our findings here suggest that metastatic competence is mainly dictated by tumor differentiation state.

A 23-gene expression signature, SCCore-GEP, captures much of the prognostic information encoded in underlying molecular features. Notably, SCCore-GEP both outperforms and complements existing clinical staging systems, suggesting that it reflects biological characteristics not captured by standard clinicopathologic measures. Based on the known functions and cellular sources of these genes, the signature primarily reflects tumor differentiation state and, to a lesser extent, the contribution of surrounding normal epithelial and immune cells. The AJCC has acknowledged that poor differentiation is associated with unfavorable outcomes, but it was considered too subjective to be included as a risk factor in a staging system^36^. By contrast, gene expression can objectively measure differentiation to complement a purely clinical/pathologic predictor.

A major strength of SCCore-GEP is that it can predict metastatic risk from biopsy material, enabling a timely risk evaluation without waiting for a full pathological assessment based on the excision. Accurately assessing metastatic risk before any therapeutic intervention may also enable risk-informed decisions regarding the optimal surgical approach (e.g., whether to perform Mohs micrographic surgery) or (neo-) adjuvant treatment. This is particularly relevant in challenging cases, such as cSCCs in frail or immunosuppressed patients and cosmetically sensitive areas. While staging systems and nomograms remain valuable tools, their performance is contingent on the availability of complete pathological data.

While our nested case-control datasets enabled the estimation of the SCCore-GEP performance at the population level, further studies are needed to identify specific patient groups that would benefit most from SCCore-GEP risk stratification. Effective cSCC patient management must also consider the risks of local recurrence, local progression, and development of multiple primaries. In summary, we generated the world’s largest population-based genomic and transcriptomic resources of cSCC, from which we developed and validated a gene expression-based signature (SCCore-GEP) with the potential to improve care of patients with cSCC.

## Methods

### Selection of tumors for molecular analyses

The tumors, selected for molecular analyses, came from a nested case-control (NCC) dataset, which was derived from a larger nationwide cohort of 19,120 patients (the D-SQUAME discovery cohort). Details of the epidemiological design, sample collection process, and tumor block slicing have been described elsewhere^18^, and the nested case-control sampling is discussed in Rentroia-Pacheco et al^20^. Briefly, through linkage of the Netherlands Cancer Registry (NCR)^37^ and the Dutch Nationwide Pathology Databank (Palga^38^), we identified all patients diagnosed with a first cSCC in the Netherlands between January 1, 2007 and December 31, 2009. Next, we retrieved their pathology reports documenting histologically confirmed primary cSCCs and metastases until 2022. Data was further linked to the Netherlands Organ Transplant Registry (NOTR^39^) to identify solid organ transplant recipients. Patients who developed metastatic disease (cases) were identified using a rule-based algorithm applied to the pathology reports^40^, followed by a manual review by two clinicians. A total of 305 cases were identified and matched to patients without metastasis (controls) using incidence density sampling, with two matching variables: pathology laboratory and metastatic risk, as assessed by the Erasmus MC clinicopathological model (EMC model^13^). The corresponding formalin-fixed paraffin-embedded (FFPE) tumor blocks were requested via the Dutch National Tissue Portal^41^. To maximize dataset completeness, pathological characteristics of all tumors were re-assessed on hematoxylin and eosin (H&E) slides by dermatopathologists blinded to both outcomes and the original pathology reports. Of the requested 305 case-control sets (N=610 primary cSCC samples), tumor blocks for 227 sets were received on time for RNA- and whole-exome sequencing.

### Whole-exome sequencing and pre-processing

A subset of the tumors selected for molecular analyses (N=174) was sequenced with whole-exome sequencing. H&E slides were macrodissected by a dermatopathologist to enrich for areas of squamous cell carcinoma. Normal skin, adjacent to the cSCC, was macrodissected and used as a source of reference, germline DNA. Genomic DNA was isolated using the QIAamp DNA FFPE Tissue Kit from 20 unstained FFPE sections of 4 and 8 µm. The DNA was sheared to an average fragment size of 300 base pairs using the Covaris E220 focused ultrasonicator. Libraries were constructed with the KAPA HyperPrep Kit (Roche, cat. no 07962363001) and enriched for exomic fragments through hybridization with KAPA HyperExome V2 probes (Roche, cat. no 9718648001). After capture, they were further amplified with 13 PCR cycles using the KAPA HyperCapture Reagent Kit (Roche, cat. no 09075828001). Samples were then paired-end sequenced (2 x 100 bp) on an Illumina NovaSeq 6000 instrument, with a target of 100M reads per sample. All the nucleic acid inputs and outputs involving library preparation, hybridization, and sequencing were quantified using Qubit (dsDNA High Sensitivity quantification), Agilent Bioanalyzer 2100 (High Sensitivity DNA run), and/or QuantStudio 5 real-time PCR system (qPCR with the KAPA Quantification Kit, Roche, cat. no 07960298001).

Fastq files were preprocessed with fastp^42^ (v.0.23.4) and aligned to the hg19 reference genome with the BWA-MEM algorithm^43^ (v.2.0.5). Duplicated reads were removed with Picard (v1.97) and base quality was recalibrated with GATK BaseRecalibrator^44^ (v.3.3.0). A total of 27 samples were excluded due to cross-contamination (N=2), mismatches between RNA-seq/WES (N=3) and insufficient power to detect somatic mutations (N=22). The last criterion was based on the percentage of callable coverage across the sequenced exome, where callable coverage corresponds to the genomic positions with sufficient power to detect a somatic mutation (STable 1, “Callable coverage (%)” column). A minimum callable coverage of 80% was required. Further details are provided in the supplementary text.

### RNA-sequencing and pre-processing

For each tumor, RNA was collected from 13 unstained sections of 4 µm FFPE tumor tissue. In contrast to the DNA isolation, the RNA was not dissected to enrich for tumor tissue – whole slide sequencing was performed. The RNA was extracted from the tissue curls using the Qiagen RNeasy FFPE kit (Qiagen, cat. no 73504). Libraries were prepared using the Illumina RNA Prep with Enrichment kit (Illumina, cat. no 20040537) combined with the Illumina Exome Panel (Cat.no 20020183), and they were sequenced on an Illumina NextSeq™ system (Illumina). Resulting RNA-sequencing data were pre-processed, aligned to the hg38 genome, and evaluated for quality control (which led to the exclusion of 12 samples). Further details pertaining to quality control exclusion criteria are provided in the Supplementary text. No clear batch effects, linked to purely technical variables, were observed, except for differences due to surgical procedure, namely, biopsy vs excision. Out of 60,295 profiled genes, 14,958 were retained for downstream analyses after filtering on transcription type, variance, and expression level.

### Somatic mutation calling

Somatic mutations (single to triple nucleotide substitutions) were identified in each sample using Mutect2^45^ (v4.2.0.0), using tumor and matched normal samples as well as the hg19 GATK panel of normals. To remove artifacts, variant calls were filtered using an orientation bias model created with the GATK *LearnReadOrientationModel* function, and variants with fewer than four supporting altered reads or an allele fraction below 4% were excluded.

Since the normal tissue was macrodissected from skin adjacent to each tumor, we explored whether tumor contamination into the normal was occurring. In approximately 5% of samples, we detected tumor contamination in the reference DNA (up to 16% of total reads). To rescue mutations potentially missed by MuTect2 due to tumor contamination, we ran Mutect2 in tumor-only mode, using a panel of normals, to rescue somatic mutations that were incorrectly filtered out. Mutations identified in this analysis were further filtered to identify variants that: (i) were not germline in 1000genomes/dbSNP, (ii) had a mutant allele fraction (MAF) in the normal <0.25, (iii) had at least 6 fold coverage in the normal sample, (iv) normal MAF was less than half of tumor MAF, and (v) tumor MAF > 0.2 ×tumor cellularity. Mutations were annotated with Funcotator (v4.2.0.0, v1.6.20190124s data source). Additionally, the TERT promoter region (chr5: 1,295,120-1,295,350) was inspected in the Integrative Genomics Viewer (IGV) in all samples, and somatic mutations showing (i) at least two reads in the normal sample, (ii) at least 2 mutated reads in the tumor sample, and (iii) tumor cellularity adjusted MAF higher than 10% were included even if they were not detected as somatic mutations by Mutect2.

Indels were identified using Pindel^46^ (v0.2.5a7). Indels were inspected in IGV and removed if the reference also had reads that misaligned in the region. This typically occurred for Indels that resided in repetitive tracks of DNA or were adjacent to germline SNPs– common causes artifacts noted in the PCAWG study^35^.

Mutations present in at least 40% of tumor cells were considered clonal. Unless specified, only these mutations were included in the analyses. Tumor mutation burden was also calculated based on clonal mutations as described by Chang et. al.^21^. Table S8 contains all somatic mutations called.

### Copy number alterations

Copy number alterations were identified using cnvkit^47^ (v0.9.6), using normal samples of each sequencing batch as panel of normals. For some sequencing batches, separate references were generated for samples with coverage above and below 100x. Chromosomal arm-level amplifications and deletions were identified based on a 0.1 cutoff applied to absolute log2 copy number ratios. Samples with tumor cellularity lower than 13.5% were excluded from copy number analyses because in these samples the cutoff of 0.1 is not sensitive enough to detect copy number deletions, as the expected absolute log2 ratio of single-copy gains would be lower than 0.1. To identify focal copy-number alterations, candidate amplifications and deep deletions were first called using CNVkit. Candidate events were then evaluated through a structured expert review process, modeled after the approach used by the Pan-Cancer Analysis of Whole Genomes working group, to assign each focal event to the most likely target gene when supported by the genomic context. Although focal copy-number alterations were uncommon in cSCC, this analysis identified additional tumors with deep deletions of CDKN2A, amplifications of CCND1, among other established cSCC driver genes.

### Tumor cellularity

We inferred tumor cellularity as previously described^21,48^. Briefly, we used the following methods, whose assumptions and formulas are detailed in the supplementary text: “Modal Somatic MAF x 2”, “Median Somatic MAF in sex chromosomes” for male patients, “Copy number deletion log ratios”, “Allelic Imbalance of SNPs over copy number neutral LOH” and “Allelic Imbalance of SNPs over copy number deletions”, as described by Chang et. al.^21^. We used the average of at least two of these methods when possible. The specific methods used for each sample are indicated in Table S1; further details are in the supplementary text. Tumor cellularity was used to compute adjusted MAFs, by dividing the unadjusted MAF by the sample tumor cellularity fraction.

### Mutational signatures

COSMIC reference mutational signatures were assigned to individual samples using SigProfilerAssignment^49^(v.0.2.3), using exome=True option and cosmic version 3.4. Samples where the signature linked to azathioprine exposure (SBS32) was detected were labeled as immunosuppressed in the whole-exome sequencing analyses.

### Nomination of driver genes

To increase statistical power, we combined the mutation calls from the 147 tumors sequenced here with those from a meta-analysis of 83 cSCCs previously performed by our group^21^.

We used IntOGen^24^ (v2024.11) to nominate cancer genes, using the default settings, but without filtering out drivers with more than 2 mutations in a sample. Genes that were identified by at least one individual tool within IntOGen and with q-value <0.01 were considered as significant. Revision of all IntOGen hits (N=27) revealed that out of 6 genes with an oncogenic role, as determined by IntOGen, 2 (*TPM3*, *SDHC*) did not have any OncoKB^25^ oncogenic mutation in the entire dataset. Additionally, a known false positive^21^, which is driven by annotation errors, affecting *KNSTRN* was called. As such, these genes were not considered as drivers. There were two genes, *LAMC1* and *IGF2BP2*, that were considered significant by IntOGen, but we did not include them as *bona fide* drivers because their mutations tended to be mono-allelic, and they do not operate in pathways known to be important in cancer or keratinocyte biology (Fig. S4C, Table S2).

Additionally, we queried OncoKB (accessed 06 October 2025), to find mutations in driver genes that are infrequently altered yet still pathogenic. Outside of IntOGen tumor suppressor driver genes, a total of 79 nonsynonymous mutations were annotated as likely gain/switch of function according to OncoKB. These were considered probably pathogenic in CSCC, except when the mutations were not in a known hotspot according to COSMIC (N=16); or have been shown to not be under selection in melanoma (N=3). TERT mutations outside C228T, C250T and CC242TT (N=18) were also not considered probably pathogenic, based on the work of Eliott et al^50^. For Loss-of-function mutations, OncoKB annotates any truncating mutations in known tumor suppressor genes as likely oncogenic. Given the high mutation burden in cSCC, we did not consider any mutation annotated as loss-of-function/likely loss-of-function as probably pathogenic in cSCC, unless they were missense mutations on a hotspot and present in COSMIC (N=2). Mutations with unknown or inconclusive effect according to OncoKB were still considered as pathogenic if they were in a clear hotspot according to cosmic (N=1). Table S3 shows all OncoKB annotated mutations in our cohort.

### Counting driver events

For each gene, all alterations labeled as probably pathogenic above were considered as driver alterations. Additionally, focal amplifications in oncogenes were considered as driver events; as were all nonsynonymous alterations and focal deletions affecting tumor suppressor genes identified under selection by IntOGen. To identify tumor suppressor genes with bi-allelic alterations or oncogenes with increased gene dosage of the oncogenic allele, we identified genes with multiple hits by identifying genes harboring more than one driver mutation, as well as genes with driver mutations showing higher than expected allele frequencies (defined as tumor cellularity adjusted MAF > 0.75). For tumor suppressor genes, we also considered the gene to have 2 hits if a chromosomal arm deletion was detected in the same tumor with a point mutation. For *USP9X,* which is located in the X chromosome, a single hit in male samples was considered equivalent to a double hit.

### Association of genomic alterations with metastatic outcome

We examined the association between each altered gene, pathway and copy number alteration with metastatic outcome, using univariate Cox proportional hazards regression models adjusted for callable coverage. For gene models, we used gene hit count as the independent variable, corresponding to the number of driver alterations in the gene, capped at 2 (i.e., 0,1 or 2). For pathway models, we grouped genes into pathways (as indicated in figure 2), summed all gene hits within the pathway and used the total hits within a pathway as the independent variable. For instance, if a tumor had bi-allelic loss-of-function mutations affecting NOTCH1 (2 hits) and an additional heterozygous mutation affecting NOTCH2 (1 hit), then the Notch pathway would have 3 total hits. For copy number alteration models, alteration presence (0 or 1) was used as the independent variable. Only genes/pathways with driver mutations present in at least 3 samples and chromosomal alterations present in at least 10 samples were assessed. The combination of pathway and chromosomal alterations that was most predictive of metastasis was identified using a multivariable regularized Cox regression model, with α=0.5 and λ chosen with cv.glmnet *lambda.1se* option. Only alterations present in at least 20% of samples were used as input features. The model was internally validated with bootstrap resampling (N=200).

### Mutual exclusivity and co-occurrence analysis

Co-occurrence and mutual exclusivity were assessed for all driver mutation pairs with driver mutations in at least 10 samples (total of 19 genes), using two-sided Fisher’s exact tests. P-values were corrected for multiple testing with the Benjamini-Hochberg correction.

### Clustering analysis

Sample clusters were found on the excision samples (N=197) using the *cola*^51^ R package (v.2.16.0) on the count data after DESeq2 variance stabilizing transformation, blinded to the metastatic outcome. Three feature selection methods were used: standard deviation, coefficient of variance and median absolute deviation, with the default clustering methods. Four solutions with K=2 showed perfect stability as measured by 1-Percentage of ambiguous clustering (PAC) (1-PAC=1), but there was intra-cluster heterogeneity in gene expression, implying that a two-cluster solution was missing some sample subtypes. As such, spherical k-means using median absolute deviation filtering with K=3 was selected as the final clustering solution because it showed high stability (1-PAC=0.953), the highest number of confident samples (i.e. samples with silhouette index higher than 0.5, N=194), and explained more outcome differences between tumors. Clusters were visualized using the 1000 genes with the highest association with the obtained sample clusters (determined using F-tests with the *get_signatures* function from the cola package). Overrepresentation analysis of each gene cluster was performed using *clusterProfiler*^52^ (v4.14.6).

Clusters found in the excision samples were validated in the biopsy samples (N=181) by projecting the biopsy samples onto the PCA space defined by the expression of the top 1000 genes in the excision samples. Clusters were then assigned to the biopsy samples using a nearest centroid classifier based on the first 3 principal components. Statistical tests used to assess the association between clinico-pathological variables/genomic alterations and genomic clusters are provided in the supplementary text.

### Differential gene expression analysis

Differential gene expression analyses between cases and controls and between immunosuppressed and immunocompetent patients were performed using the DESeq2^53^ R package (v1.46.0), with statistical adjustments for sample type (biopsy vs. excision). Gene set enrichment analysis was performed using fgsea^54^ (v1.32.4) with the *fgseaMultilevel* function. Genes were ranked by the DESeq2 Wald statistic, and enrichment was assessed against MSigDB Hallmark gene sets^55^. Pathways with an adjusted P < 0.05 were considered significant.

### SCCore-GEP development

Candidate genes for modeling were selected using a structured workflow that combined expression-based filtering, biology-informed prioritization of differentiation/progenitor-like genes, and unbiased data-driven selection of remaining genes. To select highly expressed genes for the modelling, we additionally filtered the 14,958 genes used for differential gene expression analysis down to 11,144 genes based on the average log2(TPM+1). Then, these genes were divided into two sets: (i) genes belonging to the differentiation-progenitor-like signature^31^ (*DvP* genes) (n=403); and (ii) all other genes (n=10,741), referred to as *non-DvP* genes. Within each set, another pre-filtering step was performed using DESeq2 to identify differentially expressed genes between cases and controls. This filter was performed separately within the *DvP* genes (biology-inspired feature selection) and within the *non-DvP* genes (data-driven feature selection) to promote the selection of genes involved in a diverse set of pathways.

We evaluated different combinations of modeling approaches and feature integration strategies. Specifically, we compared regularized Cox’s proportional hazards (linear model) and Random Survival Forest (RSF, a nonlinear model) models, each paired with different feature integration approaches of gene sets (no integration, early integration, or late integration at the feature or prediction level). These models were implemented as described in the Supplementary text, and only samples from complete case-control sets were used in signature development (N=366 samples). The final approach - based on Cox regression - entails two steps: 1) Cox regressions with elastic net regularization were fitted separately to the two gene sets to identify a reduced set of features associated with metastasis; then, 2) the selected features were integrated (late integration) in a single Cox regression with L2 regularization. The model was internally validated using 200 bootstrapped samples. Discriminative ability was evaluated by weighted C-index, an adaptation of the C-index adjusted to the NCC design^56^.

### SCCore-GEP genes assessment on spatial transcriptomics

Spatial transcriptomics was performed on a 10X Genomics FFPE Visium HD platform (following protocols CG000684 Rev A and CG000685 Rev B), and the resulting sequencing data were processed with the SpaceRanger pipeline (v.4.0). Supplementary Text contains further details on cluster annotation. To quantify cell compartment representations of the 23 genes included in SCCore-GEP, reads from cells assigned to each annotated cell type were aggregated per patient and expressed as a fraction of the total reads for that gene across all cells in that sample. Average proportion across patients (N=8) are reported, reflecting the relative contribution of each cell compartment to the overall expression of the signature genes.

### Validation datasets

SCCore-GEP was validated in an independent RNA-sequencing NCC dataset (N=102) derived from a second nationwide Dutch cohort of 25,921 patients, which was also part of the D-SQUAME study^18^. This dataset is a subset of the complete NCC D-SQUAME validation dataset and encompasses those samples whose sequencing data were available at the time of the study. This validation dataset differs from the discovery dataset in that patients had their first cSCC more recently (2017-2018), and no risk-matching of the controls was performed. Additionally, the SCCore-GEP was validated on an external RNA-sequencing dataset of 52 primary cSCCs^15^, where tumors were matched based on BWH staging and immunosuppression status. Limited clinical information was available for this dataset: the metastatic status was available as a binary variable (i.e., no time-to-event data), and BWH stages were extracted from samples’ annotation. Further information on how these datasets were processed is available in the supplementary text.

## Ethics statement

The study was approved by the Medical Ethics Review Committee of the Erasmus Medical Center for the discovery and validation datasets (MEC-2020-0147), as well as the scientific and privacy committees of the NCR, PALGA, and NOTR.

## Patents, Royalties, Other Intellectual Property

SCCore-GEP was patented (EP26151074, Methods for predicting the risk of metastasis in cutaneous squamous cell carcinoma (cSCC)).

## Code Availability

Code is available at https://github.com/barentroia/onco_cSCC_genomic_characterization/.

## Data Availability

Raw exome sequencing data and RNA sequencing data were made available through the European Genome Archive (Accession number EGAS50000001852). Clinical data according to the Human Tumor Atlas Network (HTAN) phase 2 data model is available upon request at the Netherlands Cancer Registry upon the following description: “HTAN dataset from data request numbers: K19.054 (D-SQUAME discovery cohort) and K23.132 (D-SQUAME validation cohort)”.

## Acknowledgements

We thank the data managers and registrars of the NCR, Palga and NOTR for compiling and linking patient and tumor data, thereby enabling tumor identification at the nationwide level. We thank the staff of the pathology laboratories for retrieving the requested tumor blocks, as well as Selin Tokez, Amber Khan, Rosa Almand, Andrya Hollatz, and Nikita Hulscher for assistance in reviewing pathology reports and processing the tumor samples. We thank Thierry van den Bosch and members of Erasmus MC Pathology Research and Trial Service (PARTS) for DNA/RNA isolation; David van Klaveren for statistical consulting and Erik Valent, Daniëlle van Keulen, Rowan Kuiper, Sheril Alex and Jvalini Dwarkasing for continuous support in the project.

## Author contributions

The project was conceptualized by LMH, MW, AHS, DoB, HvdW and BRP. Sample and clinical data collection were performed by BR-P and OFMS. Whole-exome sequencing was performed by BT, NC-P and BRP; and the resulting data was analyzed by BR-P and HS. RNA-sequencing was performed by DH and the data was analyzed by BRP, LP, JJHT, RR, YT-C and BI. Pathology revision was performed by OFMS and ALM. Spatial transcriptomics was performed by DeB and analyzed by VA and JJHT. SCCore-GEP modeling was performed by BR-P, LP, JJHT and RR. Visualization was performed by BR-P, LP, JJHT, RR and AHS. Data interpretation was performed by BRP, HS, LP, JJHT, OFMS, RR, AvH, BI, DeB, VA, CE, AB-CM, TN, HvdW, ALM, DoB, MW, AHS and LMH. The work was supervised by LMH, AHS, MW, DoB, ALM and HvdW. The original draft was written by BR-P and AHS. All authors contributed to editing and critical review of the manuscript.

---

**Supplementary Figure 1.**
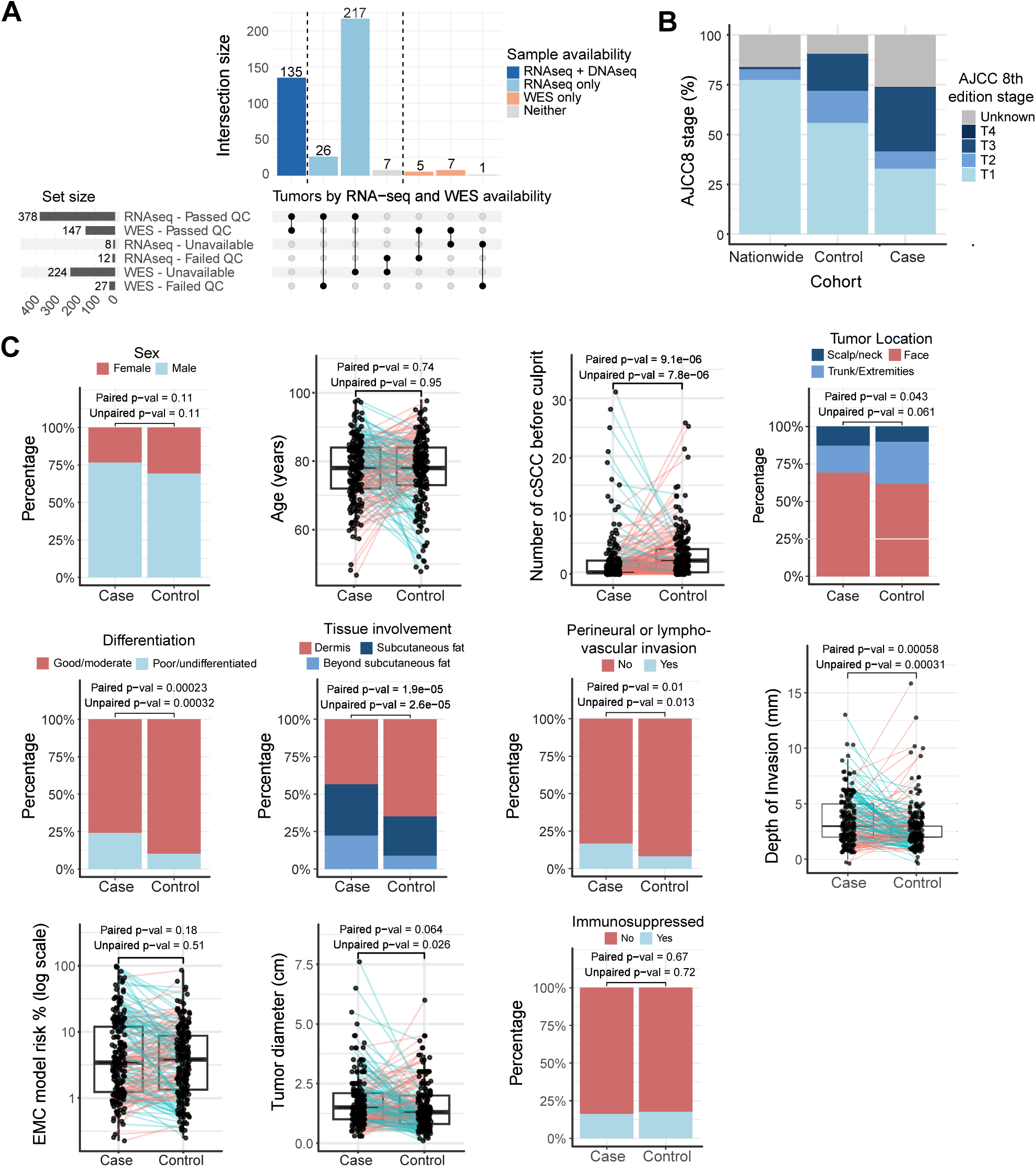
Patient and tumor characteristics of metastasizing (cases) and non-metastasizing (controls) cSCCs. **A.** Overview of available sequencing data **B.** Distribution of AJCC 8th edition stages in the nationwide cohort (N=19,120 patients), cases (N=185) and controls (N=193). As expected, the nation-wide cohort was enriched with low-risk tumors, but for this study, we chose controls that more closely matched the risk of cases. **C.** Comparison of clinical variables, tumor characteristics and the metastatic risk predicted by the clinico-pathological Erasmus MC (EMC) model. Each case was matched to a control by identifying a control from the same pathology laboratory, with longer follow-up than the case, and similar EMC risk score (see methods). As such, some clinical and histopathologic risk factors differ between cases and controls, but overall, the controls are predicted to be of similar risk. Paired p-values were computed using paired Wilcoxon and McNemar’s tests, whereas unpaired p-values were computed using Wilcoxon and chi-squared tests, for continuous and categorical variables, respectively. Lines connecting case–control pairs are colored blue when the value is higher in the case than in the respective control, and in salmon when it is lower or equal.

**Supplementary Figure 2.**
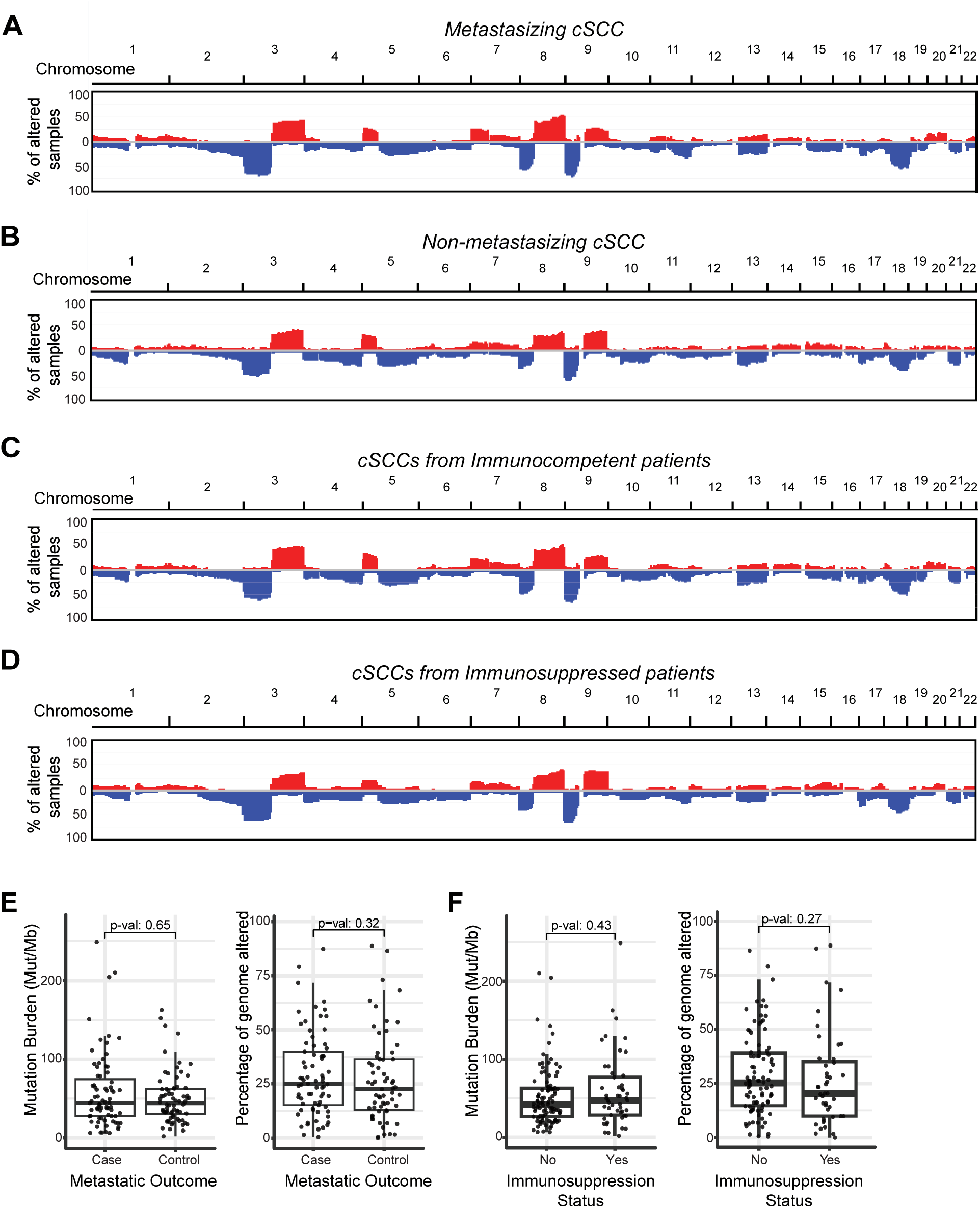
Genome-wide chromosomal alterations stratified by case/control and immunosuppression status. Data plotted as shown in figure 1C for: **A.** metastasizing cSSCs (cases); **B.** non-metastasizing cSSCs (controls); **C.** cSCCs from immunocompetent patients and **D.** cSCCs from immunosuppressed patients. See figures 3 and S6 for hazard and odds ratios indicating associations between arm-level copy number alterations and outcome/immunosuppression status. **E**. Comparison of mutation burden and percentage of the genome with a copy number alteration between metastasizing cSCC (cases) and non-metastasizing cSCC (controls), using a Wilcoxon rank test. **F.** Comparison of mutation burden and percentage of genome with a copy number alteration between immunocompetent versus immunosuppressed patients, using a Wilcoxon rank test.

**Supplementary Figure 3.**
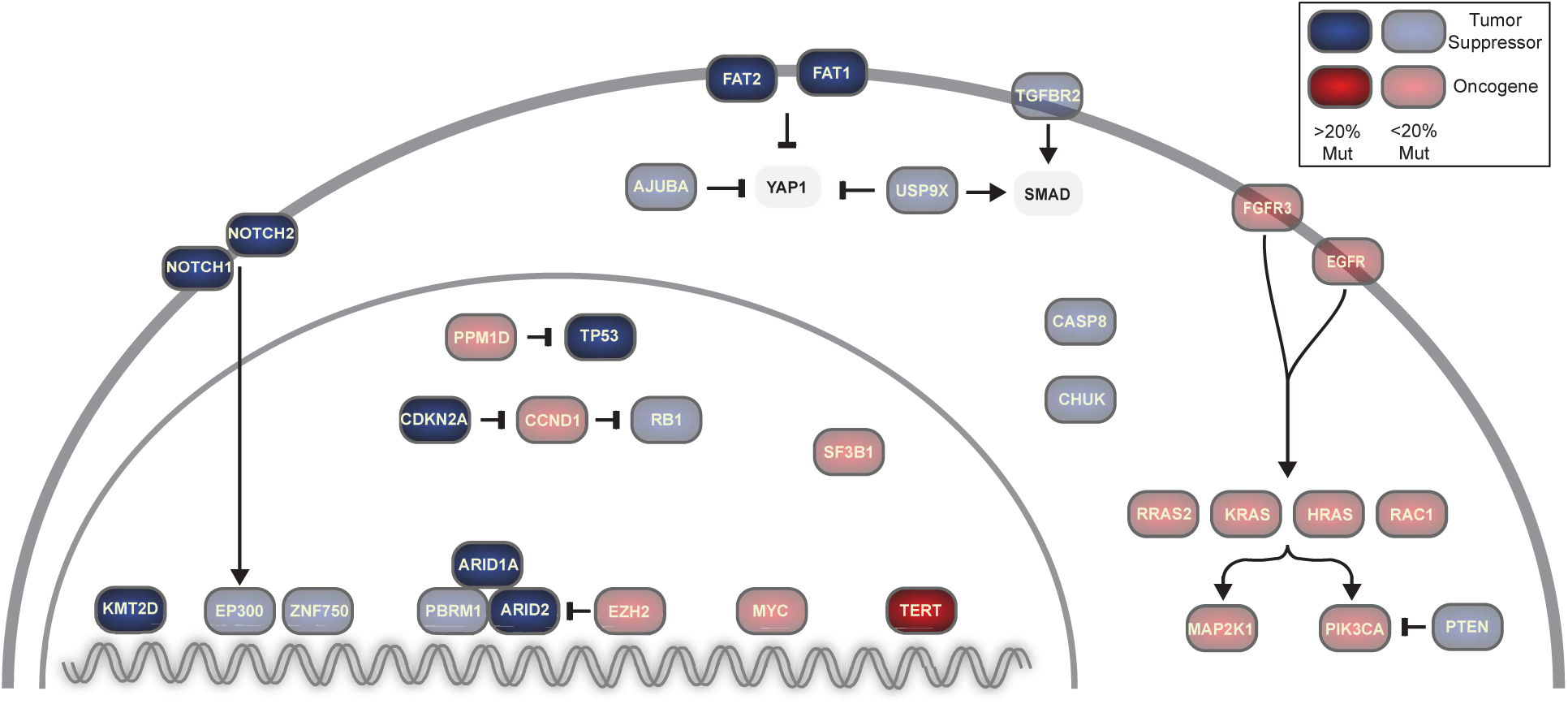
Overview of interactions among proteins encoded by driver genes in cSCC. Oncoproteins (red) and tumor suppressors (blue) with more than one underlying driver mutation (see Fig. 2A) are shown. Two additional proteins, YAP1 and SMAD (grey), are included to illustrate pathway connectivity, despite lacking evidence of selection in the present study.

**Supplementary Figure 4.**
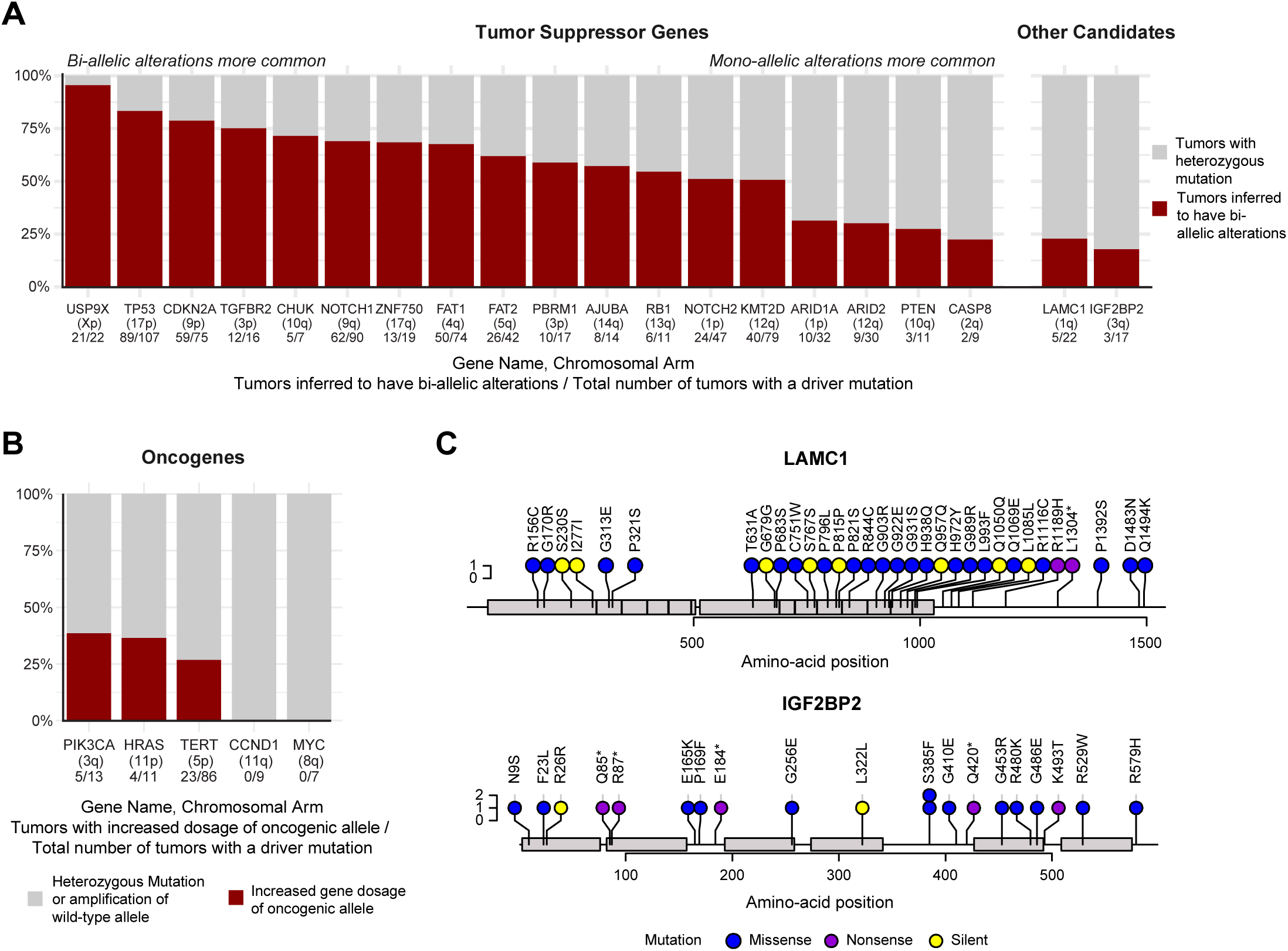
Bi-allelic alterations commonly affect tumor suppressor genes. **A.** Proportion of tumors inferred to carry bi-allelic alterations versus mono-allelic alterations in tumor suppressor genes. A tumor was inferred to have bi-allelic alterations if it had: 1. a deep deletion, 2. a point mutation and a deletion, 3. a point mutation with an elevated allele frequency (suggesting copy number neutral loss of heterozygosity), or 4. multiple nonsynonymous mutations. See methods for more details. Only genes mutated in at least 5 samples are shown. **B**. Similar to A, but reporting oncogenes with increased dosage of the oncogenic allele. **C**. Lollipop diagrams showing the distribution of mutations in two candidate genes, nominated by IntOGen, but excluded from the final gene list shown in figure 2 because their mutations were seldom bi-allelic, and their connection to cancer biology was limited.

**Supplementary Figure 5.**
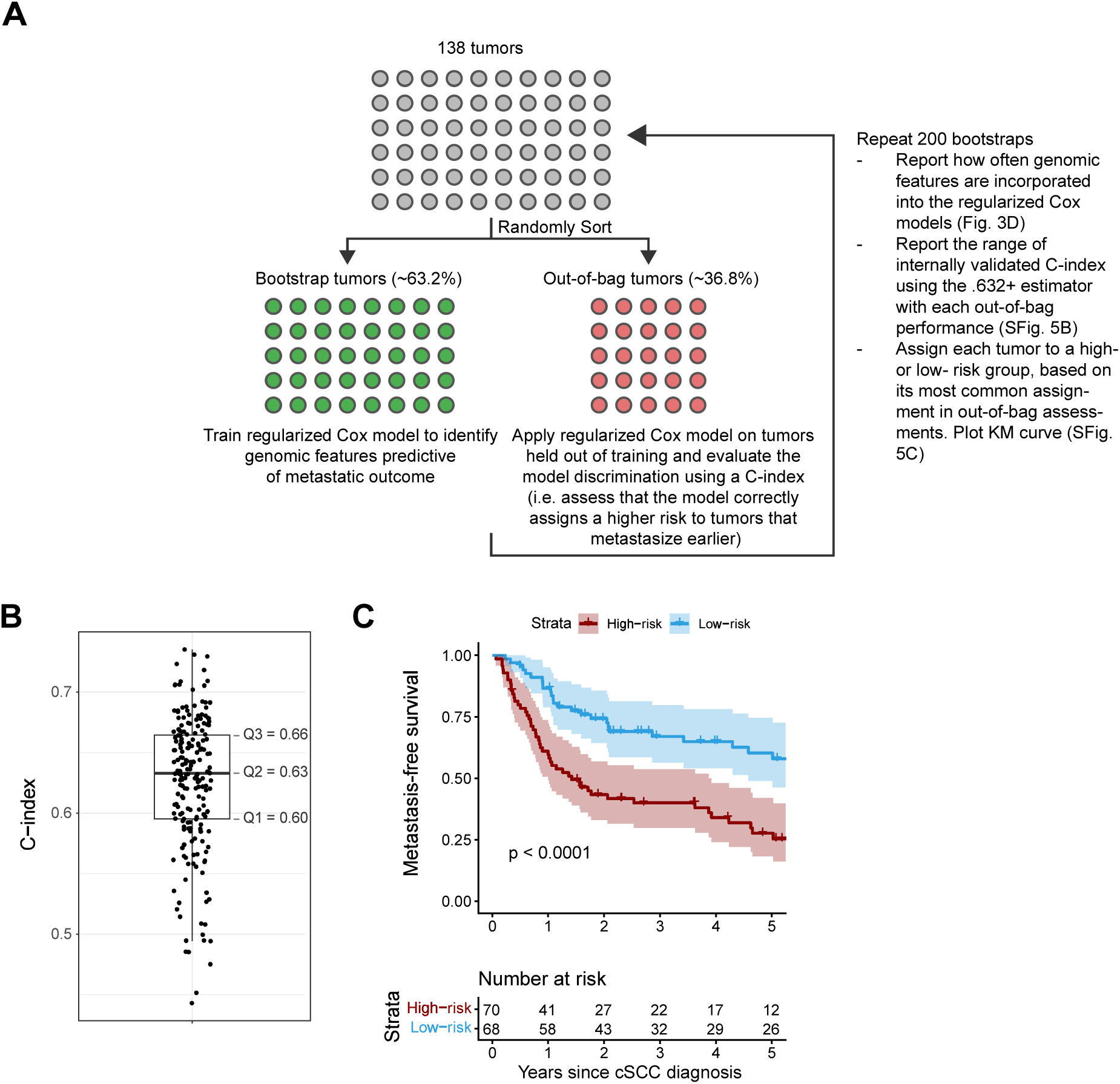
Identifying genomic features predictive of metastasis and assessing their discriminative performance. **A.** Schematic summarizing how prognostic mutational features were identified and assessed. **B.** Internally validated c-index for regularized Cox regression using pathway and copy number alterations, computed with the .632+ estimate and 200 bootstrap repetitions. **C.** Kaplan-Meier survival curves for risk groups identified using a regularized Cox regression model with pathway and copy number alterations. In each bootstrap repetition, a risk cutoff (median predicted risk) was defined on the bootstrap samples and applied to the out-of-bootstrap samples to categorize them as high or low risk. The most frequent risk label for each sample across bootstraps was considered as the final label and visualized.

**Supplementary Figure 6.**
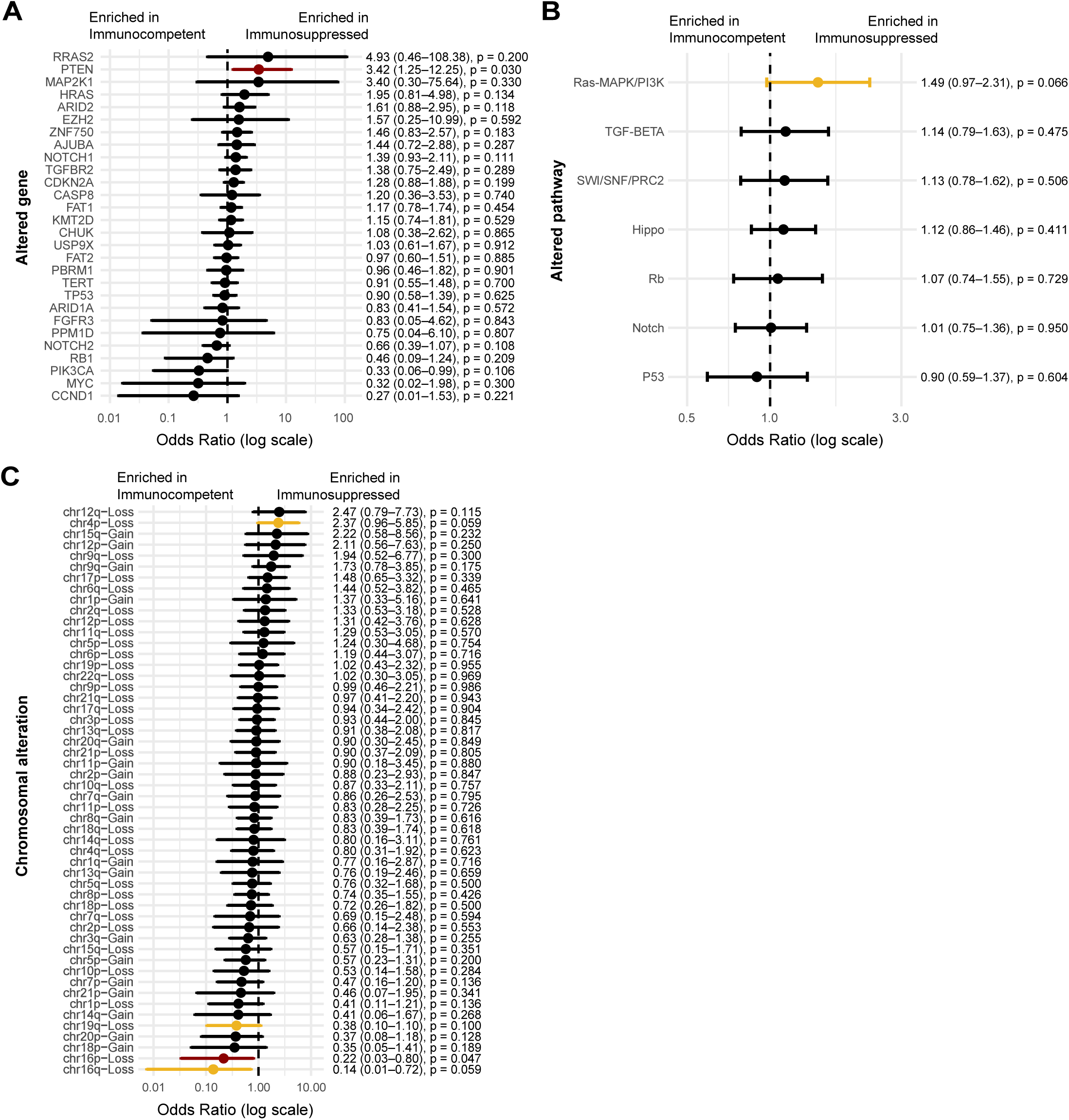
Genomic alterations associated with immunosuppression status. **A.** Association between driver alterations and immunosuppression status for all genes with driver alterations present in at least 3 samples and in both status groups. For each gene, mutation presence was coded according to the number of alterations in the gene (0,1, or 2). Association was assessed using a logistic regression model, with gene hit count as the independent variable and sample callable coverage included as an adjustment covariate. Odds ratios and Wald p-values for gene hit count are shown. p-values < 0.1 are colored in yellow, and p-values <0.05 are colored in red. **B.** Genes with driver alterations were grouped into pathways and association between the sum of all hits in a pathway and immunosuppression status was computed. Only pathways with mutations in at least 3 samples are shown. Results of logistic regression models are shown with the same settings as in A. **C**. Association between chromosomal alterations and immunosuppression status. Only chromosomal alterations present in at least 10 samples are shown. Association was assessed using a logistic regression model, with alteration presence (0 or 1) as the independent variable and sample callable coverage included as an adjustment covariate.

**Supplementary Figure 7.**
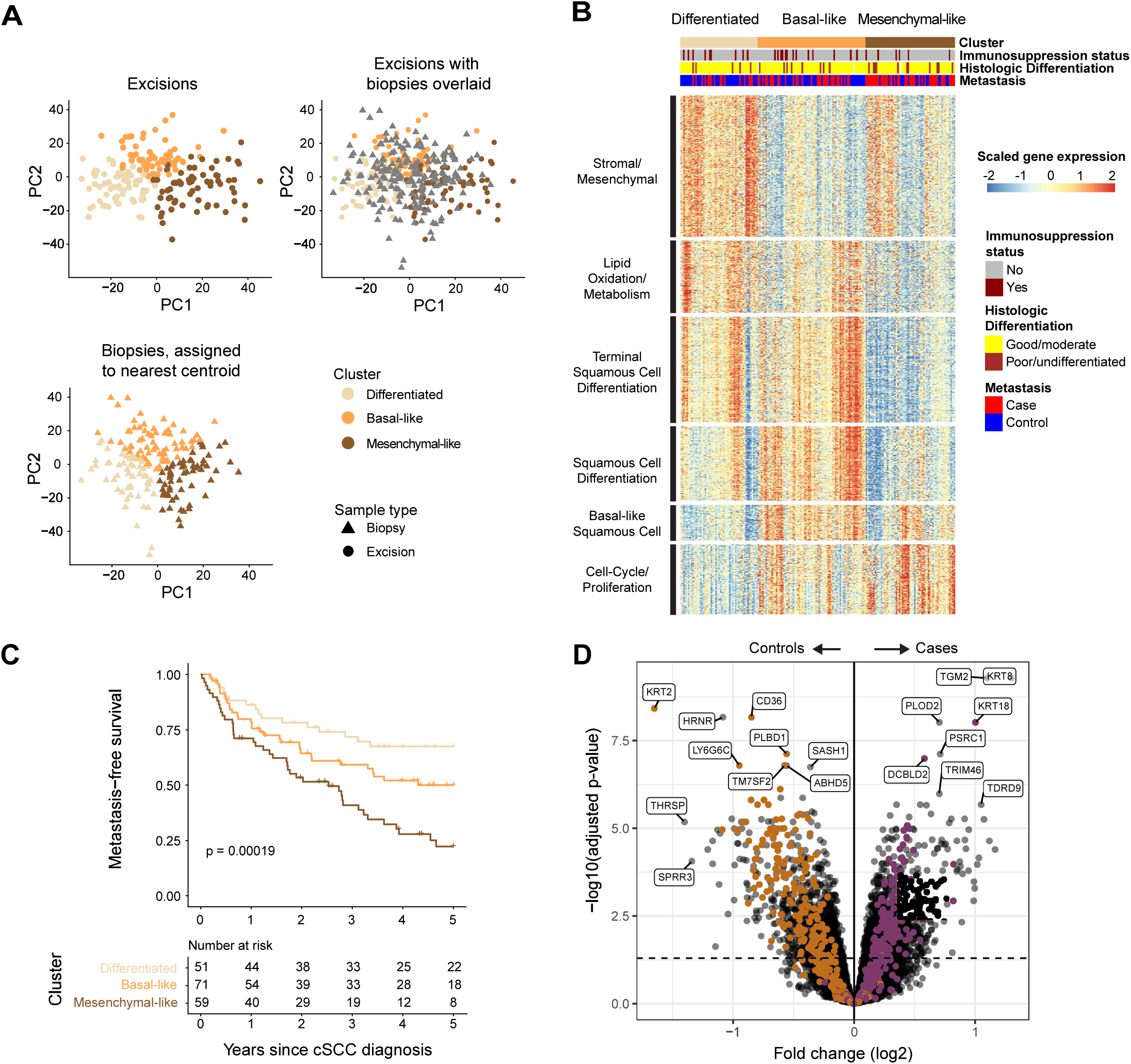
Validation of transcriptomic clusters on biopsy samples. **A**. *Top left:* Principal Component Analysis of excision samples (N=197). *Top right:* Biopsy samples (N=181) projected onto the Principal component space of the excision samples, before cluster assignement with nearest centroid. *Bottom left:* Biopsy samples after cluster assignment. **B.** Heatmap showing clustering of cSCC biopsy samples (N=181), using the top 1000 genes with the highest association to the defined clusters in cSCC excision samples. Within each cluster, samples are ordered based on hierarchical clustering using Spearman’s correlation and average linkage. **C**. Kaplan-Meier curves of the three cluster groups in the biopsy samples (N=181). Follow-up truncated at 5 years. **D.** Volcano plot showing differentially expressed genes between metastasizing and non-metastasizing CSCC, adjusted for sample type (excision vs biopsy). Significant genes were defined as those with an adjusted p-value < 0.05, indicated by a dashed horizontal line. Genes included in the Differentiation versus Progenitor (DvP) signature of Bailey et al. (252 differentiation-related and 251 progenitor-related genes) are highlighted in orange and purple, respectively.

**Supplementary Figure 8.**
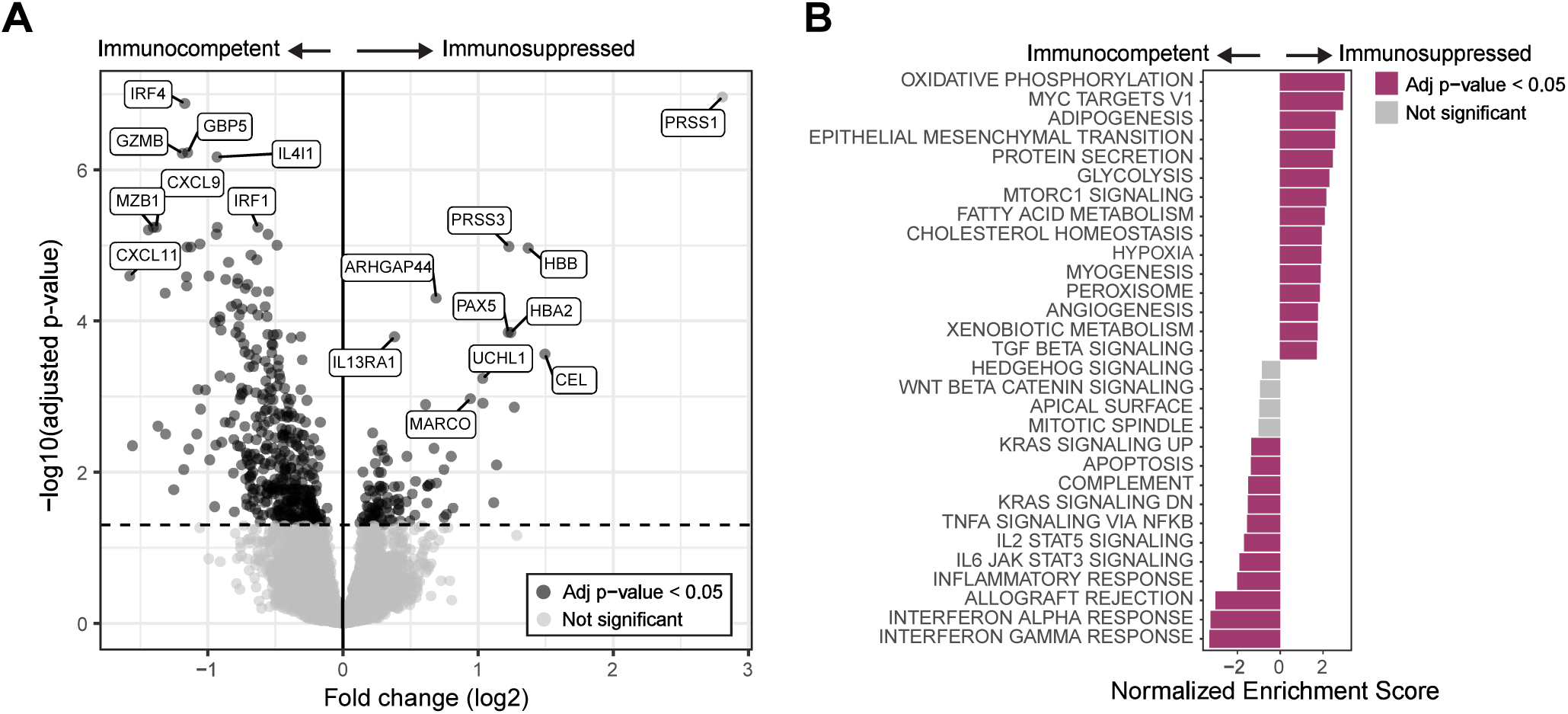
Differential gene expression analyses of cSCCs from immunocompetent versus immunosuppressed patients. **A.** Volcano plot showing differentially expressed genes between cSCCs from immuno-competent and suppressed patients, adjusted for sample type (excision vs biopsy). Significant genes were defined as those with an adjusted p-value < 0.05, indicated by a dashed horizontal line. **B.** Gene set Enrichment Analysis (GSEA) showing the top 15 upregulated and top 15 downregulated Hallmark gene sets in cSCCs from immuno-competent and suppressed patients, adjusted for sample type.

**Supplementary Figure 9.**
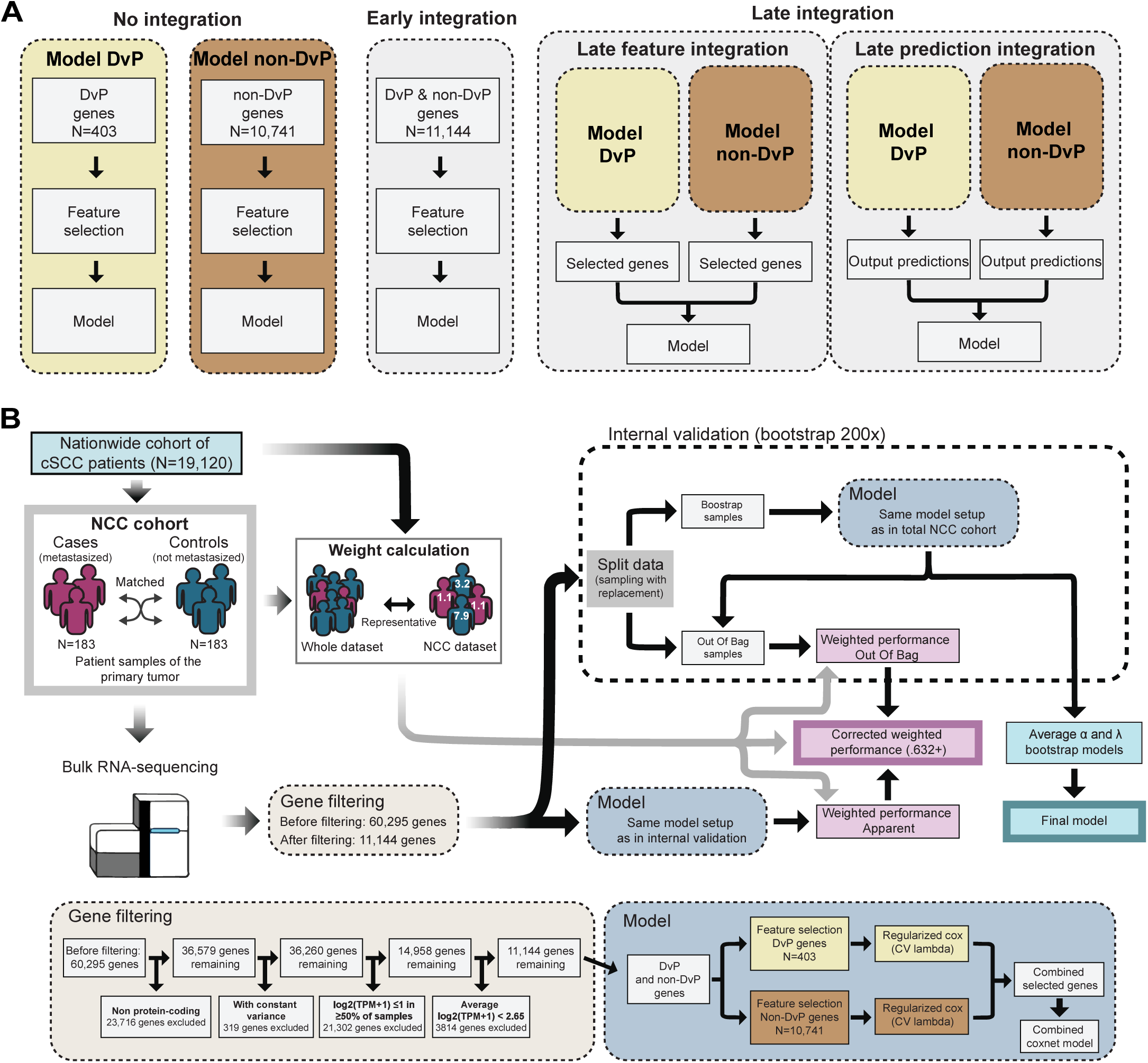
Overview of SCCore-GEP discovery. **A.** Overview of investigated input feature sets and integration strategies: no integration, early integration, and late integration. In the no integration strategy, models were built separately for Differentiation versus Progenitor (DvP) genes from Bailey et al. and non-DvP (all other genes). In the early integration strategy, a single model was built using all genes passing filtering (DvP + non-DvP). In the late integration strategy, two approaches were used: in late feature integration, separate models were built using DvP and non-DvP genes, and the selected genes were used as input for a single model; in late prediction integration, separate models were built using DvP and non-DvP genes, and the resulting predictions were used as input for a single model. **B.** Flowchart of the signature development. Discovery samples were selected using a nested case-control (NCC) study design from a Dutch nationwide cohort of 19,120 patients. Through the link to the source population, the risks estimated in the NCC apply to the general population via inverse sampling weighting. Samples were sequenced, measuring the expression of 60,295 genes. Genes with low expression or uncertain biological relevance were filtered out (gene filtering panel). The model underlying the SCCore-GEP signature was built using the filtered genes as input (model panel). Corrected weighted performance estimates were computed in a bootstrapping framework with 200 repetitions. Model hyperparameters (α and λ) averaged over all bootstrap repetitions were used for the final model. The model panel in the flowchart represents, for illustrative purposes, the feature integration scheme selected for the final model. However, if we replace it with different feature integration schemes (panel A), the flowchart can describe all modelling setups we have explored.

**Supplementary Figure 10.**
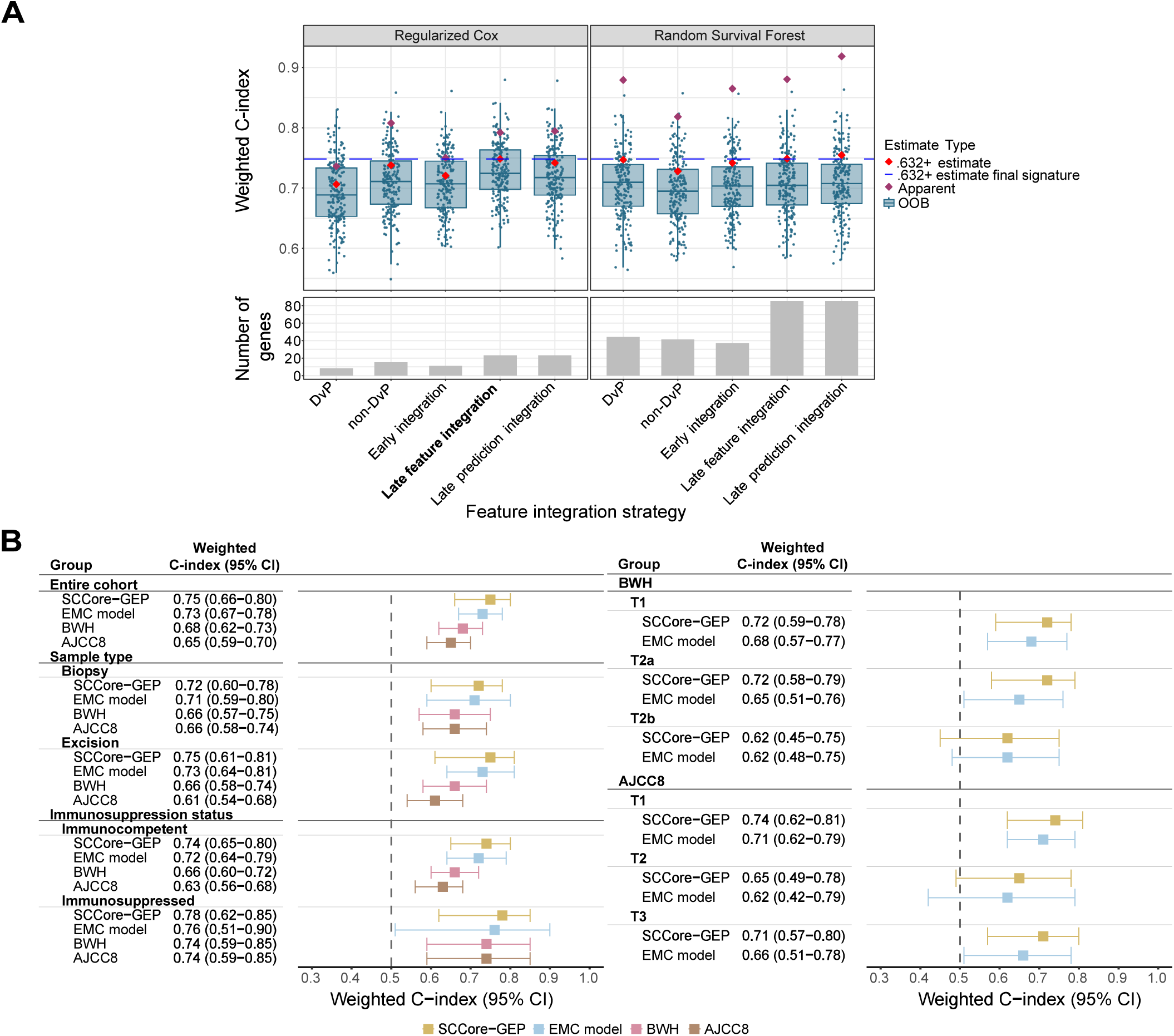
SCCore-GEP signature discovery and performance on the development dataset. **A.** Comparison of the performance of different modeling strategies. Internal validated performance (200 bootstrap repetitions) of regularized Cox and Random Survival Forest (RSF) models, each paired with 5 different feature sets, as outlined in SFig. 9A. For each feature set and model pair, the weighted C-index of the models built on the entire discovery cohort (apparent performance, indicated with a purple diamond) and their weighted C-index when applied to out-of-bag (OOB) samples (OOB performance, shown as a boxplot of 200 bootstrap repetitions) are shown. The resulting internal validation estimate of each model, assessed with the .632+ method, is indicated with a red diamond. For comparison across different modeling approaches, the .632+ estimate for the regularized Cox with late feature integration (chosen modeling approach for SCCore-GEP, in bold), is indicated by the blue dashed horizontal line. Gene numbers reflect the final signature; regularized cohort **B.** Discriminative performance of the SCCore-GEP signature in the discovery dataset. Weighted C-index and 95% confidence intervals for metastasis prediction by SCCore-GEP, EMC model, BWH, and AJCC8 staging systems in the entire discovery dataset and stratified by sample type, immunosuppression status, BWH and AJCC8 stages. Sample type refers to the type of material sequenced. Confidence intervals were obtained using 100 bootstrap samples; SCCore-GEP performance was internally validated with 200 bootstrap repetitions and bias-corrected with the .632+ method.

**Supplementary Figure 11.**
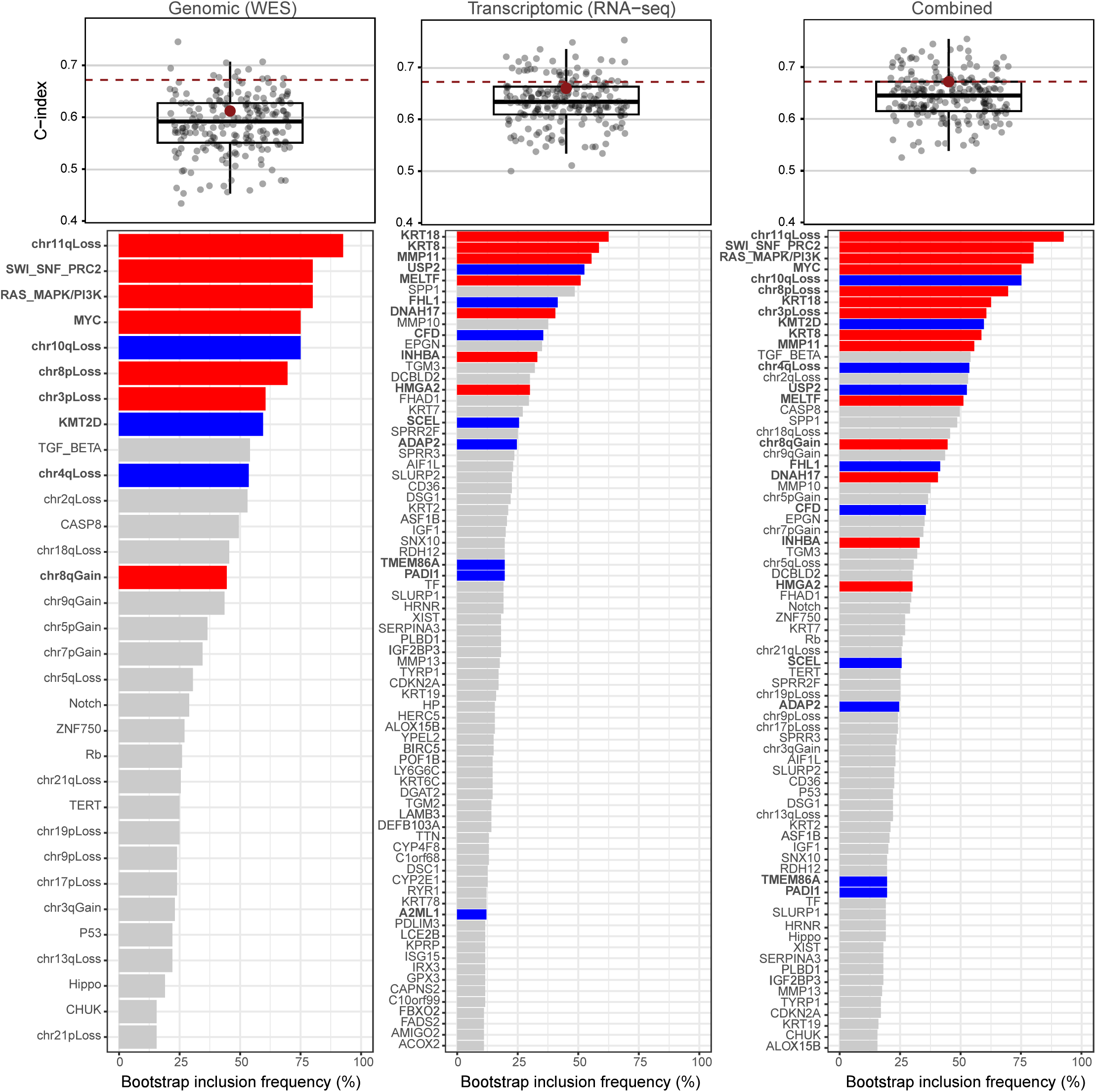
Discriminative ability and feature selection frequency of regularized Cox regression models using as input genomic alterations, gene expression features, or a combination thereof. Top: Internally validated c-indexes of regularized Cox regression models across 200 bootstrap iterations are shown for models trained using whole-exome sequencing (WES) features (left), RNA-sequencing (RNA-seq) features (middle), or a combination of both data types through late feature integration; features were selected independently from WES and RNA-seq data, followed by fitting of a Cox regression model using the selected features (right). Individual grey dots represent c-indexes obtained in the out-of-bag samples, while red dots indicate the internally validated (.632+ estimate) performance estimates. The dashed line corresponds to the internally validated performance of the combined model. WES features were prefiltered using the same prefiltering procedure as for the WES model in Fig. 3D (N = 32 prefiltered features), and RNA-seq features were prefiltered using the same prefiltering procedure as applied for the SCCore-GEP model in SFig. 9 (N = 11,142 prefiltered features). 200 bootstrap iterations are shown for WES, RNA-seq, and combined models predicting time to metastasis in the discovery cohort, with the top 75 most frequently selected features displayed per model. Colors indicate the direction of the regression coefficients; blue and red represent association with lower and higher metastatic risk respectively.

**Supplementary Figure 12.**
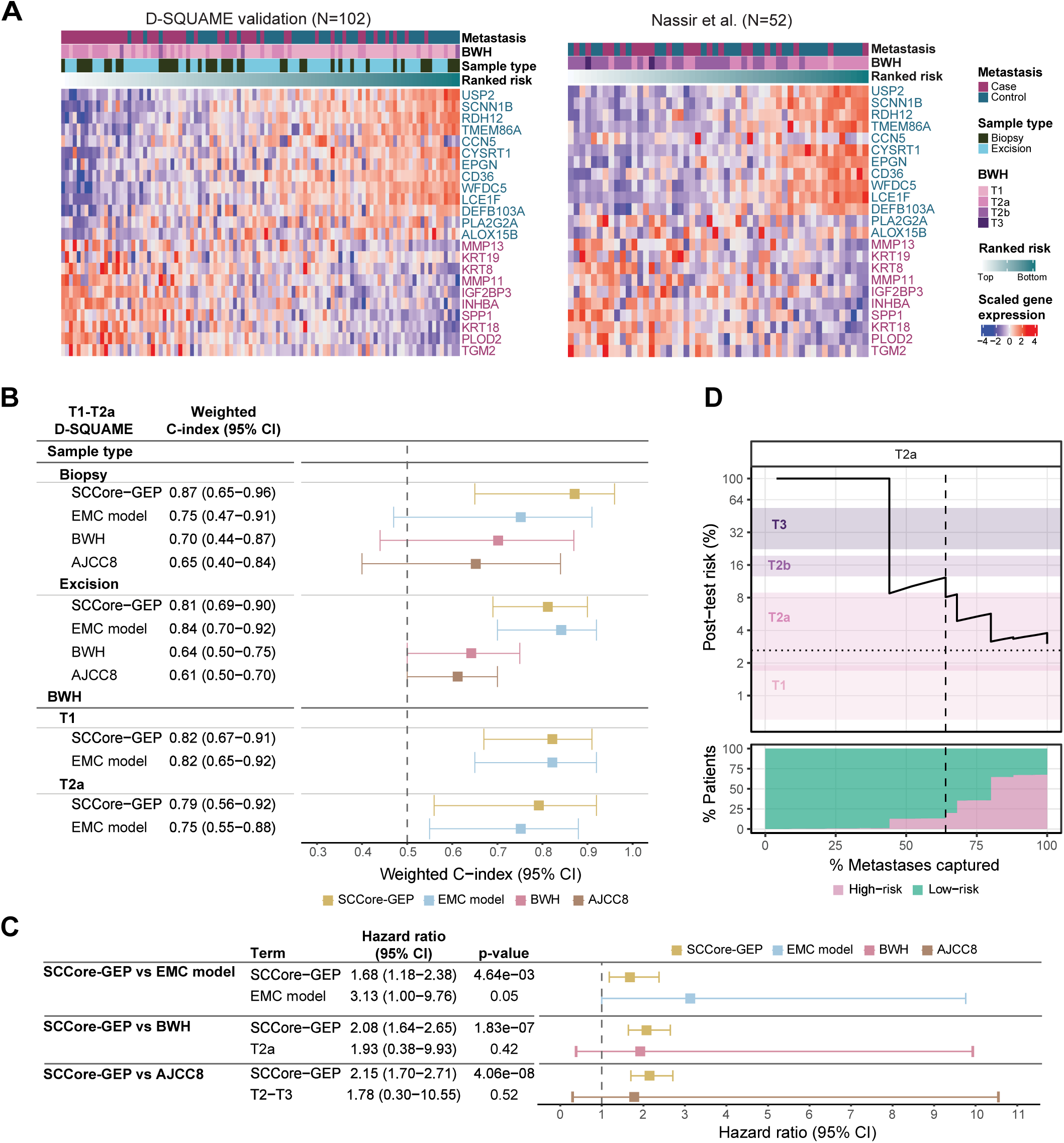
SCCore-GEP in the validation datasets. **A.** Expression of SCCore-GEP genes in bulk RNA-seq validation datasets: D-SQUAME validation (left) and Nassir et al. (right). Genes are ordered from lowest to highest SCCore-GEP coefficient; genes in blue have a negative SCCore-GEP coefficient, while genes in purple have a positive SCCore-GEP coefficient. Samples are ordered from highest to lowest SCCore-GEP risk. **B.** Assessment of the discriminative performance for metastasis prediction of the SCCore-GEP, EMC model, BWH, and AJCC8 staging systems in T1-T2a tumors of the D-SQUAME validation dataset, using weighted C-index and 95% confidence intervals. Performance is stratified by sample type (excision vs biopsy) and BWH staging. Sample type corresponds to the sequenced type of material. **C.** Multivariable analysis results of SCCore-GEP and EMC model/BWH/AJCC8, in the T1-T2a subset of the D-SQUAME validation dataset, with p-values computed using Wald tests. **D.** SCCore-GEP post-test risk probability across different risk thresholds for BWH T2a patients (N samples in NCC dataset= 36). For more details, see legend of Figure 5D.

**Supplementary Figure 13.**
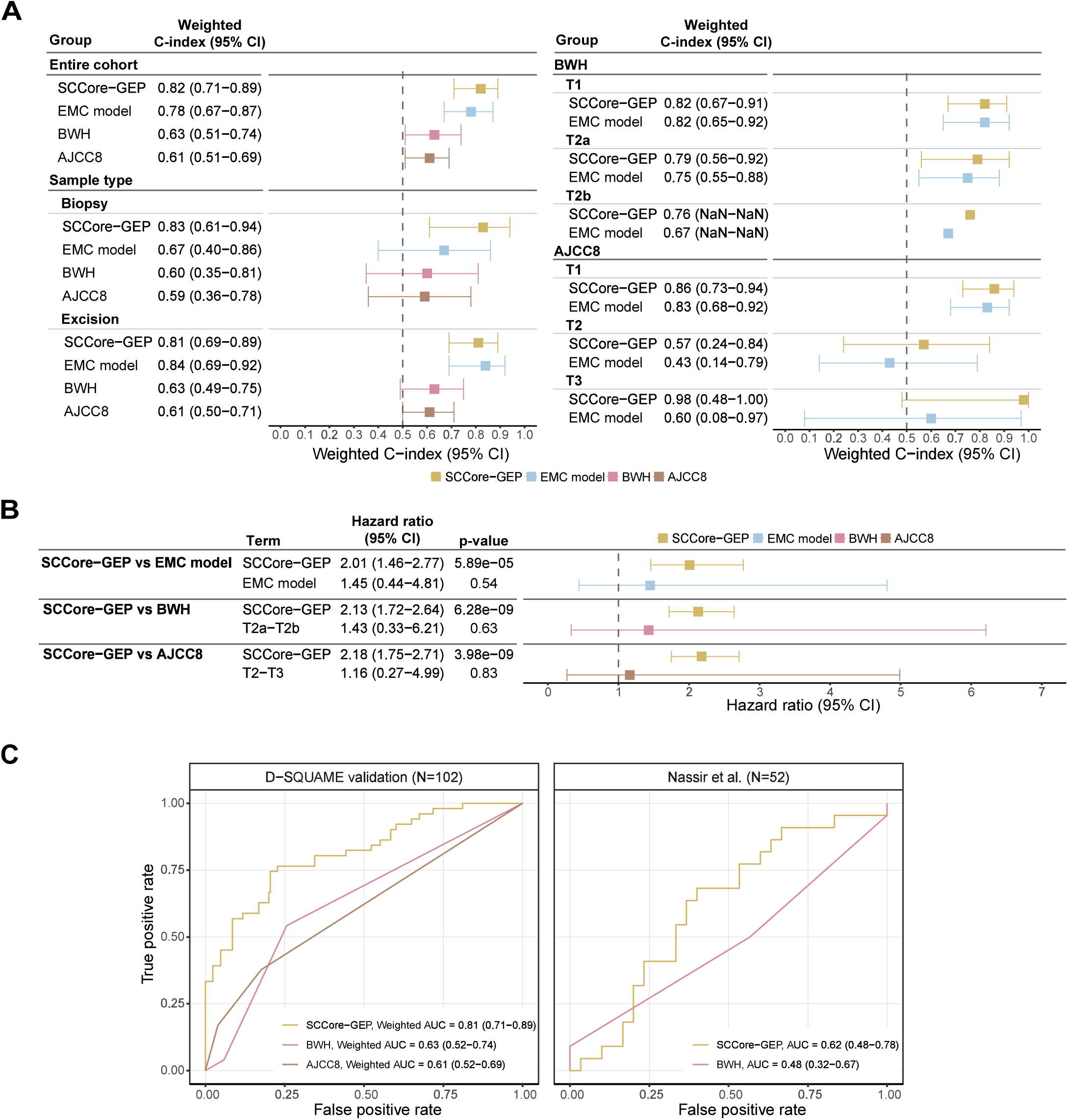
Performance of the SCCore-GEP signature on the complete D-SQUAME (N=102) and Nassir et al. (N=52) validation datasets. **A.** Weighted C-index and 95% confidence intervals for metastasis prediction of SCCore-GEP, EMC model, BWH, and AJCC8 in the D-SQUAME validation dataset and stratified by sample type, BWH and AJCC8 stages. Sample type corresponds to the sequenced type of material. **B.** Multivariable analysis results of SCCore-GEP and EMC model/BWH/AJCC8 in the D-SQUAME validation dataset, with p-values computed using Wald tests. **C.** Receiver Operating Characteristic (ROC) curves and corresponding Areas Under the Curve (AUCs) (95% CI) for metastasis prediction by SCCore-GEP, BWH, and AJCC8 in the D-SQUAME validation; and by SCCore-GEP and BWH in Nassir et al.

**Supplementary Figure 14.**
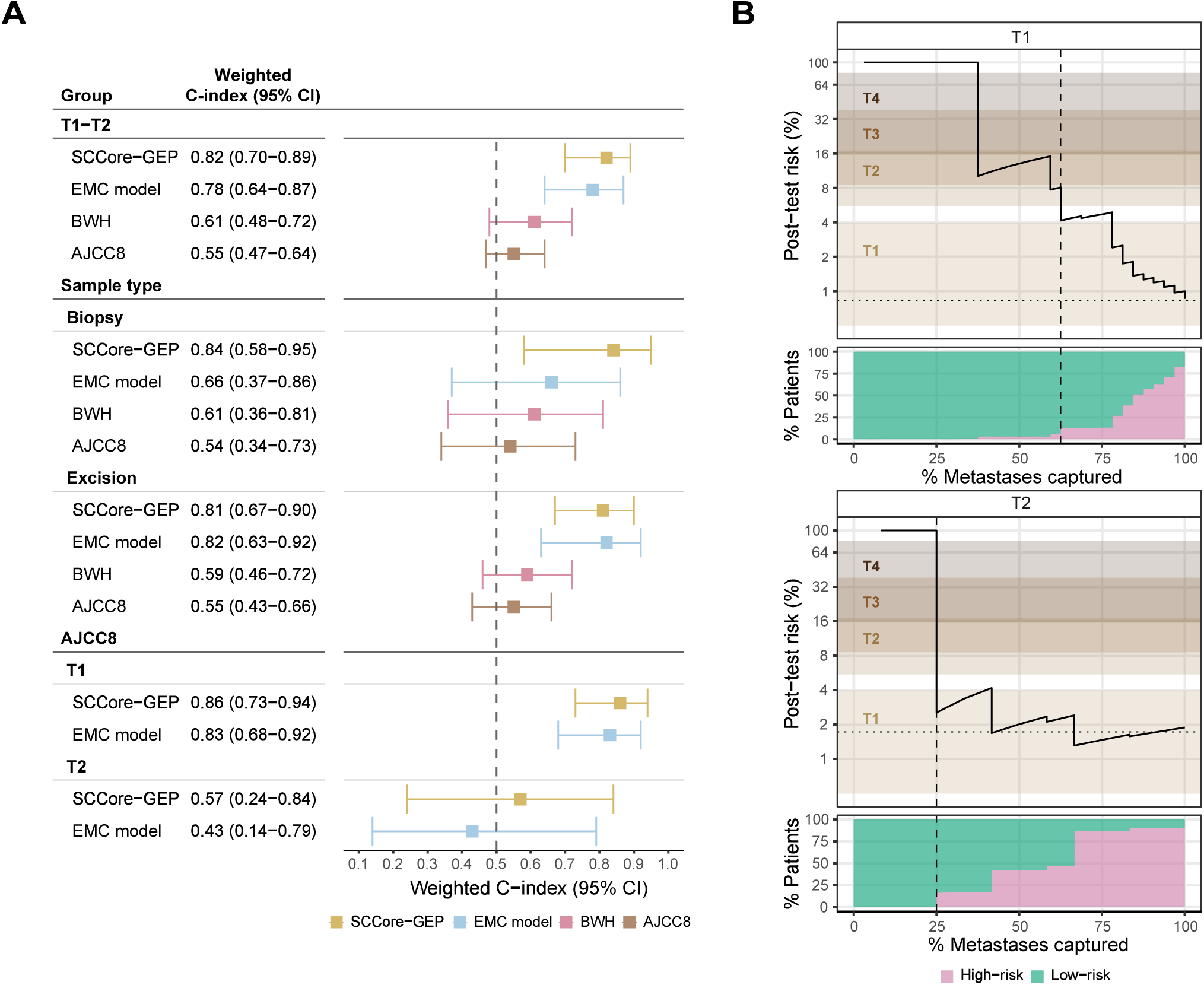
Performance of SCCore-GEP in AJCC8 T1-T2 subset of the D-SQUAME validation cohort. **A.** Weighted C-index and 95% confidence intervals for metastasis prediction of SCCore GEP, EMC model, BWH, and AJCC8 in the T1-T2 subset of the D-SQUAME validation dataset and stratified by sample type and AJCC8. Sample type corresponds to the sequenced type of material. **B.** Precision recall plot with percentage of high-risk patients in two AJCC8 patient subsets: T1 (N=75) and T2 (N=19). The upper panel shows the post-test risk (PPV/precision) versus the percentage of metastases captured (sensitivity/recall). Color bands correspond to the 95% confidence intervals of the nodal metastasis prevalence in the AJCC8 stages as reported in Zakhem et al. Horizontal dotted lines indicate the pre-test risk in the corresponding AJCC8 subset. The lower panel shows the percentage of post-test low-risk and high-risk patients versus the percentage of metastases captured (sensitivity/recall). The interval from 0 to the dashed vertical line corresponds to risk thresholds for a potential upstaging of the patients.

