## Supplementary methods for "Comprehensive molecular characterization of cutaneous squamous cell carcinoma reveals determinants of metastatic progression"

### Supplementary Information

#### *D-SQUAME discovery dataset*

##### *Sample collection*

Sample collection and processing are described in detail in Steijlen et al[1]. Briefly, out of the 305 case-control sets (N=610 samples), we were able to collect the formalin-fixed paraffin-embedded (FFPE) primary CSCC blocks of 227 case-control sets on time for sequencing. From each tumor block, we first obtained a slide of 4µm for hematoxylin and eosin (H&E) staining, followed by 27 unstained sections of 4µm: 13 for RNA sequencing, 10 for DNA sequencing, and 4 for immunohistochemistry. These sections were alternated so that tumor material for each data modality was comparable.

##### *Whole-exome sequencing*

For Whole-exome sequencing, the H&E slides were macrodissected by a dermatopathologist to identify the areas of cutaneous squamous cell carcinoma and healthy tissue. To increase DNA input, particularly from adjacent normal regions, we additionally cut a slide for H&E staining, followed by 10 consecutive slides of 8µm from an FFPE block belonging to the extremity of the excision (if available), or from the same FFPE block if only one tumor block was available. Genomic DNA was isolated from the 20 sections using the QIAamp DNA FFPE Tissue Kit. We sequenced the 174 samples in 5 batches. First, we did a pilot study with a batch of 8 samples and respective normals at GenomeScan B.V. in Leiden, The Netherlands. Sample preparation and hybridization capture for these samples was performed based on the SureSelectXT Target Enrichment System for Illumina Paired-End Sequencing Library protocol v1.8 (G7530-90000) using the Bravo Liquid Handling system. The Agilent SureSelectXT human all exon v7 capture library (5191-4006) was used. DNA sequencing was performed on an Illumina NovaSeq 6000 instrument according to manufacturer's protocols. The remaining 166 samples were prepared and sequenced at the University of California San Francisco (UCSF) in 4 batches. For each of these samples, we used all the available DNA as input up to 1000ng, and sheared it to an average fragment size of 300 base pairs using the Covaris E220 focused ultrasonicator. The treatment settings used were the following: Duty factor: 10%, Peak Incident Power: 140, Cycles per burst: 200, Duration: 80 seconds, Mode: Frequency sweeping, Temperature: 6oC. Libraries were constructed with the KAPA HyperPrep Kit (Roche, KK8504), using the low DNA input protocol. In brief, the fragmented DNA samples were end-repaired, ligated to IDT8 or IDT10 dual index adaptors, and amplified. Equal amounts, by mass, of uniquely indexed DNA sample libraries were mixed with COT DNA in a 1.5 mL Eppendorf DNA LoBind Tube. The tube was left uncapped and covered with a sterile, non-woven rayon gas exchange membrane. The multiplexed sample library pools were dried in a Thermo Scientific Savant DNA120 SpeedVac Concentrator at a HIGH drying rate (65oC), resuspended in 65 uL of PCRgrade water, and processed for capture. This was followed by exome enrichment through hybridization with KAPA HyperExome V2 probes (9718648001) and post-capture amplified with 13 PCR cycles using the KAPA HyperCapture Reagent Kit (09075828001). Samples were then paired-end sequenced (2 x 100 bp) on an Illumina NovaSeq 6000 instrument, with a target of 100M reads per sample. Samples were sequenced in 5 batches. All the nucleic acid inputs and outputs involving library preparation, hybridization, and sequencing were quantified using Qubit (dsDNA High Sensitivity quantification), Agilent Bioanalyzer 2100 (High Sensitivity DNA run), and/or QuantStudio 5 real-time PCR system (qPCR with the KAPA Quantification Kit, Roche, KK4854).

*RNA-sequencing*

For RNA-sequencing, total RNA was extracted using the RNeasy FFPE kit from Qiagen (Qiagen, Hilden, Germany, cat. no. 73504) following the manufacturer's protocols, with paraffin removal performed with the deparaffinization solution (Qiagen, cat. no. 19093). Concentration and purity of the RNA samples were measured by a Nanodrop 2000 (Thermo Fisher Scientific, Waltham, MA, USA) spectrophotometer. Library preparation with exome capture followed by enrichment step was performed with the Illumina RNA Prep with Enrichment kit (Illumina, Inc, San Diego, CA, USA, cat. no. 20040537) according to the manufacturer's instructions for FFPE material. Sequencing was performed on an Illumina NextSeq™ platform, using the NextSeq™ 500/550 High Output Kit v2.5 (Illumina, cat. no 20024906). Out of the 227 sets, 32 case-control sets were excluded due to poor RNA yield (N=1) and low pre-library concentration (N=31); the remaining 195 sets proceeded to sequencing.

*WES data pre-processing and quality control*

Fastq files were pre-processed as described in the main text. To verify sample integrity, correspondence between tumor and normal pairs, and between RNA-seq and WES samples of the same patient was checked with NGSCheckmate version 1.0.1, after liftover of RNAseq bam files to hg19 using picard LiftoverVcf v3.1.1. Two samples were discarded due to cross contamination of tumor normal pairs and three samples due to mismatch between RNA-seq and WES data. All other samples did not show any mismatch. Sex identified with copy number analyses was compared to reported sex on the clinical records and no mismatches were found.

Tumor cellularity was computed using multiple methods: "Modal somatic MAF x 2", "Median Somatic MAF in sex chromosomes" for male patients, "Copy number deletion log ratios", "Allelic Imbalance of SNPs over copy number neutral LOH" and "Allelic Imbalance of SNPs over copy number deletions". All methods are described in [2,3], except for the method based on copy number deletion log ratios. In this method, we inferred tumor cellularity based on the observed log2 copy number ratio ($c$) for a deletion in a clonal and diploid sample. For deletions in such samples, normal cells present two copies of the genomic segment, whereas the tumor cells present only one. Consequently, $c$ takes the form:

$$c = log_2(\frac{x + 2(1 - x)}{2x + 2(1 - x)})$$

where $x$ corresponds to the tumor cellularity in the sample.

Power to detect somatic mutations was assessed in each tumor-normal pair, as described previously[2]. Briefly, for normal samples, a 6x fold coverage was targeted to capture both alleles at each genomic position. For tumor samples, a minimum 8x effective coverage (coverage × tumor cellularity) was targeted, which would correspond to an expectation of 4 mutant reads at a genomic mutation, assuming equal sampling of each allele. The callable genomic footprint was defined the number of bases where a somatic mutation could be detected according to these criteria and was calculated using footprints[4] (v.0.1.0). For each sample, the smallest of the tumor or normal callable footprints was considered. Samples for which less than 80% of the sequenced exome (N=22) did not reach these thresholds were excluded.

Tumor in normal contamination was estimated for each sample by computing the average of the mutant allele frequency in the normal sample across all clonal somatic mutations identified in the corresponding tumor sample, and multiplying this value by two.

*RNA-sequencing data pre-processing and quality control*

To obtain counts and $\log_2$(TPM+1) gene expression matrices for the discovery dataset samples, an in-house bioinformatics pipeline was run. BCL files were converted to FASTQ files and demultiplexed (bcl2fastq2 (v2.20), Illumina), and run statistics were generated from the interop metric file. Adapter and read quality trimming were performed, followed by read mapping to obtain gene expression values and additional quality metrics. Sequencing adapters and parts of reads with a Q-score below 20 were trimmed (TrimGalore v0.6.6[5]; cutadapt v3.4[6]; seqtk v1.3[7]). The trimmed reads were used for downstream processing. FastQC (v0.11.9)[8] was run to obtain QC metrics (e.g., GC content, number of reads, and Q-scores). Reads were aligned against the reference genome by a two-pass STAR run (STAR v2.7.10a)[9], resulting in binary sequence alignment (BAM) files, from which a summarization of read location (coding, intronic, untranslated regions (UTR), intergenic, ribosomal) and normalized transcript coverage was created by the Picard/GATK tool CollectRnaSeqMetrics (GATK v4.2.0.0[10]; Picard v2.25.0[11]). Post-alignment processing was performed using samtools/htslib (v1.12)[12]. Next, transcript-level abundances (counts and transcripts per million (TPM)) were quantified from FASTQ files using Salmon (v1.9.0)[13] against a reference transcriptome generated from the UCSC Golden Path hg38 reference genome (accessed on November 23, 2023) using the GENCODE v38 gene annotation. Transcript-level abundances were then merged into gene-level expression values using the tximport R package (v4.4.0)[14].

After the preprocessing, samples were excluded if they failed in (1) the percentage of coding and percentage of mapped/aligned reads in Salmon and STAR, or (2) more than 3 QC criteria points. Failing criteria were defined as follows: Q30-scores<80, GC content>60, percentage of coding<65%, percentage of rRNA>20%, percentage of mapped/aligned reads by Salmon<75%, and percentage of mapped/aligned reads by STAR<75%. For the calculation of the number of failed QC criteria, percentage of coding, percentage of rRNA, and percentage of mapped/aligned reads in Salmon and STAR were counted as 0.5 points. Additionally, samples were excluded when they appeared as clear outliers in the coverage over normalized genes or in the PCA plots. In total, 12 samples were excluded based on the QC criteria mentioned above, resulting in 378 samples for transcriptomic analyses. Specifically for SCCore-GEP signature development, samples matched to the excluded samples (N=12) were additionally excluded, resulting in 366 perfectly paired samples (N=183 case-control sets).

Out of 60,295 genes measured, initial exploratory analyses were restricted to protein-coding genes and long non-coding RNAs (N excluded=23,716). Additionally, genes with constant variance (N excluded=319) and low expression, where low expression was defined as genes with $\log_2$(TPM+1) below 1 in at least half of the samples (N excluded=21,302), were filtered out. The remaining 14,958 genes were used for downstream analysis.

*Assessment of statistical associations between genomic clusters and clinico-pathological variables or genomic alterations*

The association between genomic clusters and clinico-pathological variables was assessed using the following statistical tests: Chi-squared tests for categorical variables with high frequencies, Fisher's exact test for categorical variables with low frequencies, and Kruskal-Wallis test for continuous variables.

The association between genomic clusters and number of gene/pathway hits was assessed using Poisson regressions. The association between genomic clusters and genomic summaries (Tumor mutation burden, fraction of genome altered, percentage of UV mutations or tumor cellularity) was assessed using linear regressions. In both cases, percentage of callable coverage was included as a covariate.

***D-SQUAME validation dataset: sample and data processing***

Sample collection of the D-SQUAME validation dataset followed a similar protocol as described in Steijlen et al[1]. The RNA-sequencing data was generated using a protocol similar to the one used for the discovery dataset. Deviations from the discovery protocol were as follows: sequencing was performed on NextSeq™ 2000, instead of on NextSeq™ 550. And for sequencing, the NextSeq™ 2000 P3 XLEAP-SBS™ reagent kit (Illumina, cat. no. 20100989) was used instead of the NextSeq™ 500/550 High Output Kit v2.5.

Raw BCL files of the available samples (N=102) were pre-processed into gene expression data as described for the discovery dataset. The following tool versions differed from those previously reported, and additional tools were used for QC metrics:

STAR (v2.7.9a)[9], Salmon (v1.10.1)[13], FastQC (v0.12.1)[8], TrimGalore (v0.6.10)[5], cutadapt (v4.9)[6], Picard (v3.0.0)[11] and samtools (v1.16.1)[12].

The same QC criteria used on the discovery dataset were applied to the D-SQUAME validation dataset. All samples passed the QC criteria, resulting in 102 samples for downstream analysis. The gene filtering was also the same as described for the discovery dataset.

No batch correction was required when applying the SCCore-GEP on the validation dataset, because Principal Component Analysis showed no systematic differences between D-SQUAME discovery and validation samples.

***Nassir et al. dataset: sample and data processing***

Raw FASTQ files of 75 samples belonging to the Nassir et al. dataset[15] were downloaded from the NCBI Gene Expression Omnibus (GEO) data repository (GSE284467), and pre-processed to gene expression data as described for the discovery dataset. The following tools versions differed from those previously reported, and additional tools were used for QC metrics: STAR (v2.7.9a)[9], Salmon (v1.10.1)[13], FastQC (v0.12.1)[8], TrimGalore (v0.6.10)[5], cutadapt (v4.9)[6], Picard (v3.0.0)[11] and samtools (v1.16.1)[12].

The same QC criteria used on the discovery cohort were applied to the Nassir et al.[15] dataset. This resulted in the exclusion of two samples, leaving 22 samples with metastasizing CSCC, 30 samples with non-metastasizing CSCC, and 21 normal tissue samples (designated as "control" in Nassir et al.[15]). Genes were also filtered based on the gene filtering criteria defined for the discovery dataset. Of the 60,293 genes measured, 47,523 were excluded, leaving 12,770 for downstream analysis.

Principal Component Analysis with the D-SQUAME discovery and Nassir samples showed clear separation between the two datasets. To correct for the difference in distribution between the gene expression of the two datasets and thus validate the SCCore-GEP signature, batch correction with *ComBat* (*sva* R package, v3.54.0[16]) was performed. The *ComBat* function was applied, taking the discovery dataset as the reference batch, and metastasis status and BWH staging as covariates in the model matrix.

Extensive clinical data were not available in the GSE284467 repository, but BWH staging could be derived from the sample names. Samples without BWH staging annotation were assumed to be stage T2b (N=16). The assumption was based on the comparison between the patients' characteristics retrieved from the sample names annotation and Table S1 in Nassir et al.[15]: there were 24 T2b patients in Table S1 in Nassir et al.[15], whereas there were only 10 T2b samples based on the available annotation. Metastatic status was available in the GEO repository metadata, whereas no time-to-event data was reported.

***Spatial transcriptomics***

Eight CSCC samples from the discovery dataset were selected for spatial transcriptomics (4 case-control pairs: 3 T1, 4 T2a, and 1 T3 according to BWH stages). Spatial transcriptomics was performed on a 10X FFPE Visium HD platform (following protocols CG000684 Rev A and CG000685 Rev B). Square areas of 6.5x6.5 mm were selected from the H&E slide, including both tumour and normal areas. The resulting sequencing data were processed with the 10x Genomics SpaceRanger pipeline (v.4.0). Cell clusters were annotated using an ontology-integrated framework for cell-type enrichment. The reference database was constructed by integrating 14 public and curated sources, including CellMarkerDB, PanglaoDB, and CellxGene, which were standardized using Cell Ontology (CL) and Uberon tissue identifiers to ensure nomenclature consistency. Cluster-specific marker genes were tested for enrichment against this master database using a hypergeometric test, and p-values were corrected using a global Benjamini-Hochberg False Discovery Rate (FDR) across all clusters. A minimum overlap of at least two genes ($k >= 2$) was required for an annotation to be considered. Final cell-type assignments were determined by integrating the hypergeometric enrichment score and the significance level (FDR).

To quantify cell compartment representations of the 23 genes included in SCCore-GEP, reads from cells assigned to each annotated cell type were aggregated per patient and expressed as a fraction of the total reads for that gene across all cells in that sample. Average proportion across patients (n=8) are reported, reflecting the relative contribution of each cell compartment to the overall expression of the signature genes.

##### *Multiple imputation*

In the D-SQUAME discovery and validation datasets, some clinicopathological variables required for staging and computing the EMC clinicopathological model risk were missing in some patients (Table S1). Namely, tumour location and tumour diameter were sometimes not reported in the pathology reports, and depth of invasion, tissue involvement, perineural invasion (PNI), lymph vascular invasion (LVI), and bone invasion could not be determined from the available H&E slide of the tumour in some patients. Therefore, we imputed these missing values using the procedure described below. The imputation procedure was very similar in both the discovery and D-SQUAME validation datasets, and small differences between them are explicitly mentioned.

For bone invasion, PNI or LVI, which correspond to binary variables: presence = “Yes” or absence = “No”, values were imputed to “No”. This is because we assumed these variables were not missing at random: their absence from the report likely reflects their absence in the tumour, as they would otherwise most likely have been reported. For the remaining variables, values were assumed to be missing at random and were imputed using *multiple imputation* by chained equations (*mice* R package v3.17.0[17]). Twenty-five imputed datasets were created, using the following variables as predictors: sample type, metastatic status, time to metastasis or end of follow-up or death (whatever comes first), vital status at the end of follow-up and corresponding follow-up time, age, sex, organ transplant receiver status or presence of haematological malignancies (in the D-SQUAME validation cohort, these two variables were combined into a single variable "immunosuppression status"), number of prior CSCC, tumour location, tumour diameter, depth of invasion, Breslow thickness, tissue involvement, resection margin, PNI, PNI or LVI, solar elastosis, peritumoral infiltration, BWH, AJCC8, EMC model risk. Default *mice* imputation methods and 40 iterations were used. Model convergence was assessed through visual inspection of the convergence plots.

To reduce computational costs and simplify visualization, imputed BWH stages were aggregated to compute and plot threshold-based performance metrics in the D-SQUAME validation dataset (Fig. 5D & SFig. 12D and 14B), and to visualize BWH stages associated with each sample in the D-SQUAME discovery and validation datasets (Fig. 5A & SFig. 12A). Namely, for both D-SQUAME discovery and validation datasets, the twenty-five imputed datasets were merged into a single one by taking the median and mode of continuous and categorical variables, respectively.

Weighted C-index and weighted Area under the Receiver Operating Curve (AUC) in the D-SQUAME validation dataset from the twenty-five imputed datasets were pooled using the *pool_auc* function in the *psfmi* R package (v1.0.0)[18].

##### ***Computation of sampling weights in the D-SQUAME discovery and validation datasets***

To ensure that performance metrics estimated in NCC datasets are comparable to those that would be obtained in the source population, metrics must be weighted using inverse sampling weights, as explained in Rentroia-Pacheco et al[19]. Briefly, inverse sampling weights are assigned to every individual in the NCC dataset to account for the in-built bias when sampling the NCC dataset. In other words, the weights compensate for the differences in composition between the NCC dataset and the source population. In our setting, the composition bias is due to different proportions of cases and controls, as well as the use of matching variables. In the discovery dataset, the matching variables were pathology lab, type of first procedure (biopsy or excision), and metastatic risk as assessed by the EMC clinicopathological model[20]. In the D-SQUAME validation dataset, the matching variables were the pathology lab and the type of procedure. Therefore, the discriminative ability of the model was evaluated using a weighted C-index and/or a weighted AUC, which are respectively adaptations of the C-index and AUC to the NCC design[21]. Weights were computed with inverse probability weighting[22], where the weight of each individual $(w_i)$ corresponds to the inverse of the probability $(p_i)$ of that individual being sampled from the source population ($w_i = \frac{1}{p_i}$).

In an ideal NCC study design, sampling probabilities of cases are equal to 1 as all cases are sampled. However, in practice, it might not be feasible to sample all cases[23]. Specifically, in our study, we did not receive all metastasizing CSCC samples that were requested from the national database, and some samples failed during RNA-sequencing; thus, the sampling probabilities of the cases were adjusted accordingly, by dividing the number of cases in the NCC dataset ($N_{cases,NCC}$) by the number of cases in the source population ($N_{cases,source}$): $p_{case,i} = \frac{N_{cases,NCC}}{N_{cases,source}}$.

Sampling probabilities of the controls were estimated using the logistic regression estimator[22,24]. Briefly, the sampling probability of each control was obtained after fitting a logistic regression model to the dataset comprising all patients in the nationwide dataset, except for cases. In this model, the outcome variable indicates whether a control is included in the NCC dataset or not, and the covariates are the matching variables. It is computed as follows:

$$p_{control,i} = P(S_i|x_i) = \frac{1}{1 + \exp[-(\alpha + \boldsymbol{\beta}^{\mathsf{T}}\boldsymbol{x}_i)]}$$

Where:

- $S_i$ is the outcome variable: it takes the value 1 if the control $i$ was included in the NCC dataset, and 0 otherwise.
- $\boldsymbol{x}_i$ is the covariate (matching variables) vector for control $i$
- $\alpha$ is the model intercept
- $\boldsymbol{\beta}$ is the vector of regression coefficients of the logistic regression model, containing the covariates' coefficients.

Often, CSCC patients develop more than one primary CSCC. Because each tumour can serve as a potential control, patients with more primary CSCCs have a higher chance of being sampled. To account for this, we calculated the sampling probability of each control as the complement of the probability that none of the tumours of the control were sampled. Specifically, we (1) calculated the sampling probability of each tumour (i.e. tumour-specific sampling probability), (2) computed the corresponding probability that each tumour was not sampled (i.e. tumour-specific probability of not being sampled), by taking the complement of each tumour-specific sampling probability, and (3) multiplied these probabilities across all tumours of a given patient to obtain the probability that the patient was not sampled. Of note, this assumes that sampling was performed with replacement, which is a safe approximation given the high number of controls.

In the calculation of the weights, the pathology lab was not taken into account, as some pairs were rematched (i.e., samples are not matched based on the pathology lab anymore), and considering the pathology lab as a matching variable resulted in extreme weights in the case of samples coming from pathology labs that sent only a few samples. However, we have checked that the Spearman's correlation between weights with and without adjustment for the pathology lab was high (>0.9).

In addition, when computing discovery dataset weights, missing values in variables needed to compute the metastatic risk in the source population were imputed as follows:

- categorical variables (tumour location, differentiation, sample type): mode in the source population;
- numerical variables (tumour diameter): median value in the source population.

##### ***Model development***

The model development pipeline is depicted in the signature discovery flowchart (SFig. 9). It consists of two main building blocks: gene filtering and model building. Gene filtering is performed, as a first step, on the entire dataset, whereas model building is performed within a bootstrap-based internal validation scheme. We explored different model types (Cox model and random survival forests), and different feature integration schemes (no integration, early integration, late feature or late prediction integration; as depicted in SFig. 9A). However, the model building block, as depicted in SFig. 9B, reflects the final settings used for SCCore-GEP development (Cox model and late feature integration).

*Gene filtering*

Gene filtering aims at removing noisy genes. The same filtering criteria as those described in the "*D-SQUAME discovery dataset: RNA-sequencing data pre-processing and quality control"* section were applied, with the additional exclusion of 3,814 genes with mean $\log_2$(TPM+1) below 2.65. This threshold corresponds to the 20$^{th}$ percentile of the DESeq2-normalized counts, and the corresponding $\log_2$(TPM+1) value was determined by fitting a linear regression model mapping $\log_2$(TPM+1) to DESeq2-normalized counts. After this filtering step, 11,144 genes remained for model development. Of note, these filtering criteria were applied to the entire discovery cohort, since they do not use any information regarding the model outcome. To increase robustness to outliers, gene expression values (TPM) were log-transformed ($\log_2$(TPM+1)).

*Model building*

Model building entails an initial (heuristic) feature selection, followed by model fitting. Feature selection was performed using DESeq2[25] on the input counts matrix, with metastatic status as the outcome of interest. To favour the identification of genes associated with metastasis independently of sample type and sex, these variables were included as covariates in the DESeq2 design matrix. To identify robust features leading to large expression changes, genes were filtered based on their significance and fold-change between metastasizing and non-metastasizing CSCC as follows: first, only genes with an adjusted p-value lower than 0.05 and an absolute $\log_2$(fold change) greater than 0.5 were retained; second, if more than 50 genes met these criteria, the $log_2$(fold change) threshold was iteratively increased in steps of 0.1 until the number of remaining genes was below 50. Filtering genes by the largest absolute $log_2$(fold change) and limiting the maximum number of input genes to 50 reduced overfitting compared to filtering by adjusted p-value only. For the model fitting step, we systematically investigated different feature integration approaches (SFig. 9A) in combination with two different models, regularized Cox regression and Random Survival Forest (*RSF*).

*Feature integration*

Model building was performed taking as input two different feature sets: genes belonging to the differentiation-progenitor-like signature (*DvP* genes), defined by Bailey et al.[26] (n=403); and all other genes (n=10,741) (*non-DvP* genes). To explore how the information from *DvP* and *non-DvP* genes could be best combined to improve model performance, we investigated four feature integration strategies: no integration, early integration, late feature integration, and late prediction integration (SFig. 9A). In no integration, we build separate models for the *DvP* genes and non-*DvP* genes. In early integration, all genes (DvP + non-DvP) were used as input for feature selection using DESeq2, and the selected genes were used to fit a single model. In late feature integration, feature selection with DESeq2 and model fitting were applied separately to each feature set. Successively, the features used by the individual models were combined to fit a single model (in the case of Cox regression, the model combining the two sets of features used L2 regularization). In late prediction integration, the predictions from the individual models were combined and used as input to a single model.

*Regularized Cox*

Regularized Cox regressions (Elastic net and Ridge) were implemented using the *glmnet* R package (v4.1-8)[27,28]. Elastic net uses both L1 and L2 penalties, promoting sparse models through the L1 penalty, and adequately dealing with correlated input features through the L2 penalty. Ridge regression uses the L2 penalty only and therefore does not enforce sparsity. In both models, the regularization parameter λ was chosen as the value whose corresponding model showed a cross-validated error within one standard error of the minimum error (*lambda.1se* option, implemented with the *cv.glmnet* function). The α parameter, which controls the balance between L1 and L2 penalties in Elastic net, was selected by grid search between 0.6 and 1 (with a step of 0.05), to maximize the training performance (measured in terms of weighted C-index).

*Random Survival Forest*

Random Survival Forests were implemented using the *randomForestSRC* R package (v3.4.0)[29]. Models were trained using the *rfsrc* function, and hyperparameter tuning was performed using *tune.rfsrc*. Default parameters were used, except for the following modifications to control model complexity and reduce overfitting: (1) the number of genes used at each split was set to the square root of the total number of predictor genes, with a minimum of one gene; (2) the minimum number of samples allowed in a terminal node was increased to a range of 20 and 30; and (3) tree depth was limited to a maximum of six, which prevented high apparent performance (>90%); lower depth values decreased out-of-bag performance without noticeably reducing apparent performance.

*Internal validation*

The apparent weighted C-index ($wCindex_{app}$) was obtained by performing the filter-based feature selection and model fitting to the entire discovery cohort, followed by applying the resulting model to the entire cohort. To obtain internally validated weighted C-indexes, internal validation was done using bootstrap[30] (N repetitions = 200). Briefly, in each repetition, samples were drawn with replacement from the discovery cohort, resulting in a bootstrap sample, whereas samples not included in the bootstrap sample were part of the out-of-bag (OOB) sample. The entire model-building procedure (including filter-based feature selection) was performed on each bootstrap sample. Each bootstrap model was then applied to the corresponding OOB sample, obtaining the OOB weighted C-index ($wCindex_{oob}$). Bias-corrected weighted C-indexes were computed using Efron's .632+ estimator[31] as follows:

$$Efron's\ .632+\ wCindex = (1-\alpha) \times wCindex_{app} + \alpha \times wCindex_{oob};$$

where

$$B = 200;$$

$$wCindex_{oob} = \frac{1}{B}\sum_{b=1}^{B} wCindex_{b,\,oob}\,;$$

$$R = \frac{wCindex_{oob} - wCindex_{app}}{\gamma - wCindex_{app}};$$

$$\gamma = no\ informantion\ rate = 0.5$$

$$\alpha = \frac{0.632}{1 - R \times 0.368}$$

To calculate confidence intervals, an outer bootstrap[32,33] loop was implemented and repeated 100 times. For each outer bootstrap, samples drawn with replacement were given as input to the inner bootstrap, and the bias-corrected weighted C-index was estimated. 95% confidence interval lower and upper bounds were respectively computed as the 2.5th and 97.5th percentiles of the 100 bias-corrected weighted C-indexes.

*Final gene signature*

Regularized Cox regression, with late feature integration, was chosen as the final signature. Among the investigated modelling approaches (SFig. 10A), we chose the one that resulted in the highest out-of-bag weighted C-index (implying less overfitting) both in the entire cohort and within BWH T-stages, while featuring a low number of genes (N=23).

For the final model, the entire model building procedure was applied on the discovery dataset, and the Cox regressions with input *DvP* or *non-DvP* genes as well as the Cox regression integrating the two sets of features were fitted using the average alpha (or alpha = 0, for the Cox regression integrating the two sets of features) and average lambda of the corresponding Cox regression models across the 200 bootstrap repetitions (this selection of lambda can be considered as a form of stability selection[34]).

##### *Evaluation of model performance in validation cohorts*

The performance of the final model was evaluated in two validation cohorts: D-SQUAME validation and Nassir et al.'s [15] datasets. First, both datasets were transformed to match the data used in the model development ($\log_2(TPM+1)$), and then the model was applied to compute predictions for each sample. In the Nassir et al.[15] dataset, due to a lack of time-to-event data, the model performance was evaluated in terms of Area Under the Curve (AUC). In the D-SQUAME validation cohort, the model performance was evaluated both in terms of weighted AUC and weighted C-index. The 95% confidence intervals were computed using bootstrap[32,33]. For performances in the D-SQUAME validation dataset, a bootstrap was done by keeping matched samples together during the sampling procedure, and lower and upper values were respectively computed as the 2.5$^{th}$ and 97.5$^{th}$ percentiles of the distribution of performances. For performances in the Nassir et al.[15] dataset, as well as for stratified performances in the D-SQUAME validation dataset, where not all samples have their corresponding pair in the same strata, 95% confidence intervals were computed using the *boot* and *boot.ci* functions with percentile method (*boot* R package v1.3-28.1[35,36]). In both cases, the number of bootstrap repetitions was set to 100.

In both validations, the model performance was estimated in the entire datasets as well as within BWH T1-T2a and AJCC8 T1-T2 patients. Additionally, in the D-SQUAME validation dataset, the model was evaluated within the following subsets: two different sample types (Biopsy and Excision) and within different BWH and AJCC8 stages. Evaluation within AJCC8 stages was not possible in the Nassir et al.[15] dataset, as this information was not available.

##### *Multivariable analysis*

To assess whether the SCCore-GEP is independent of BWH staging, AJCC8 staging, or the EMC model, multivariable survival analyses were performed using weighted Cox regression models (*survival* R package v3.8-3[37,38]), taking as input SCCore-GEP risk predictions, together with BWH stages, AJCC8 stages, or EMC model risk predictions, respectively.

The SCCore-GEP risk predictions (where the risk indicates the risk of developing metastasis within 5 years) were obtained by performing Cox recalibration, where the weighted Cox regression was fitted taking as input the logarithm of the negative logarithm of SCCore-GEP predictions.

In the multivariable analysis, the following transformations were applied before fitting weighted Cox regression models:

- logit-transformation of SCCore-GEP risk predictions;
- logit-transformation of EMC model risk predictions;
- AJCC8 T2-T3 samples were merged into a single subset, due to the limited number of T2 and T3 samples;
- BWH T2a-T2b samples were merged into a single subset, due to the limited number of T2b samples (only for multivariable analysis in entire D-SQUAME validation cohort).

Pooled log(HR) and corresponding 95% confidence intervals were obtained using Rubin's rules[39], where standard errors were estimated through bootstrap (number of bootstrap repetitions = 100). Wald test p-values (null hypothesis: log(HR) = 0) were computed using pooled estimates[39], calculated as described above, and Barnard-Rubin degrees of freedom[40].

Multivariable analyses were performed both in the BWH T1-T2a subset of the D-SQUAME validation cohort, as well as in the entire D-SQUAME validation cohort.

##### *Calculation of threshold-based performance metrics*

Threshold-based performance metrics (i.e., sensitivity, Positive Predictive Value (PPV), % high-risk) plotted in Fig. 5D and SFig. 12D and 14B are weighted adaptations of threshold-based metrics for binary data[21], used to estimate performances in the source population. Metrics were computed at each unique prediction score, leading to N=62 decision thresholds in BWH T1, N=36 in BWH T2a, N=75 in AJCC8 T1 and N=19 in AJCC8 T2.
